# VarLOCK - sequencing independent, rapid detection of SARS-CoV-2 variants of concern for point-of-care testing, qPCR pipelines and national wastewater surveillance

**DOI:** 10.1101/2022.01.06.21268555

**Authors:** Xinsheng Nan, Sven Hoehn, Patrick Hardinge, Shrinivas N Dighe, John Ukeri, Darius Pease, Joshua Griffin, Jessica I Warrington, Zack Saud, Emma Hottinger, Gordon Webster, Davey Jones, Peter Kille, Andrew Weightman, Richard Stanton, Oliver K Castell, James A.H. Murray, Tomasz P Jurkowski

## Abstract

The COVID-19 pandemic continues to pose a threat to the general population. The ongoing vaccination programs provide protection to individuals and facilitate the opening of society and a return to normality. However, emergent and existing SARS-CoV-2 variants capable of evading the immune system endanger the efficacy of the vaccination strategy. To preserve the efficacy of SARS-CoV-2 vaccination globally, aggressive and effective surveillance for known and emerging SARS-CoV-2 Variants of Concern (VOC) is required. Rapid and specific molecular diagnostics can provide speed and coverage advantages compared to genomic sequencing alone, benefitting the public health response and facilitating VOC containment. In this work, we expand the recently developed SARS-CoV-2 CRISPR-Cas detection technology (SHERLOCK) to allow rapid and sensitive discrimination of VOCs, that can be used at point of care and/or implemented in the pipelines of small or large testing facilities, and even determine proportion of VOCs in pooled population-level wastewater samples. This technology aims to complement the ongoing sequencing efforts to allow facile and, crucially, rapid identification of individuals infected with VOCs to help break infection chains. Here, we show the optimisation of our VarLOCK assays (<u>Var</u>iant-specific SHER<u>LOCK</u>) for multiple specific mutations in the S gene of SARS-CoV-2 and validation with samples from the Cardiff University Testing Service. We also show the applicability of VarLOCK to national wastewater surveillance of SARS-CoV-2 variants. In addition, we show the rapid adaptability of the technique for new and emerging VOCs such as Omicron.

**Short abstract:** The COVID-19 pandemic continues to pose a threat as emergent and existing SARS-CoV-2 variants endanger the efficacy of the vaccination strategy. Rapid surveillance for known and emerging SARS-CoV-2 Variants of Concern (VOC) would be assisted by effective molecular diagnostics procedures. Here we develop the recent SARS-CoV-2 CRISPR-Cas detection technology (SHERLOCK) for rapid and sensitive discrimination of VOCs to complement sequencing and allow rapid identification of individuals infected with VOC. We show our assays can be implemented with test samples in the pipelines of large testing facilities, as well as determine the proportion of VOCs in pooled population level wastewater samples and has potential applicability at point of care. We demonstrate the optimisation of new VarLOCK assays (<u>Var</u>iant-specific SHER<u>LOCK</u>) for multiple specific mutations in the S gene of SARS-CoV-2 and validate these with samples from the Cardiff University Testing Service, as well as the applicability of VarLOCK to national-level wastewater surveillance of SARS-CoV-2 variants. We also demonstrate the rapid adaptability of the technique for new and emerging VOCs such as Omicron.

## Introduction

Genetic variations in the SARS-CoV-2 genome emerge during viral replication. Whilst mutations are often inconsequential, several mutations have already emerged conferring phenotypic advantages to the virus and posing additional challenges for national and global COVID-19 responses. To date the World Health Organisation (WHO) has assigned five Variants of Concern (VOC) labelled: Alpha – B.1.1.7, Beta – B.1.351, Gamma – P.1, Delta - B.1.617.2 and most recently Omicron – B.1.1.529. These variants display increased transmissibility (Alpha < Delta < Omicron), increased risk of severe illness and hospitalisation (Alpha) and raise concerns about immune response evasion (Beta, Delta, Omicron), posing increased risks of re-infection and vaccine escape combined with increased transmissibility. Such variants pose a risk of immune escape and their surveillance have been a longstanding point of concern.

Genomic sequencing has been extremely successful in identifying new variants, in the tracking of viral lineages to understand viral introduction events and transmission[1], as well as monitoring virus evolution during the pandemic in near real-time. Genomic sequencing remains the gold-standard for tracking viral mutations and identifying emerging Variants of Concern. Once specific mutations of VOCs are known, which may be identified anywhere in the world, sequencing becomes a powerful indicator of local prevalence, but it is also a lagging indicator due to the high resource requirements and potential delays in sequencing. In practice, this currently means results take several days following a PCR positive SARS-CoV-2 test and only a sub-set of positive patient samples can be sequenced, corresponding to typically ~10% of positive identified cases in the UK, but varies geographically. This delay and sub-sampling impact the speed and efficacy of the public health response as it is highly likely that once known VOCs are picked up through genomic sequencing, there will already be multiple other undetected cases in the community.

There is therefore a requirement for combining genetic virus surveillance by sequencing with sequence-specific molecular diagnostics for rapid discrimination of variants in either laboratory or point of care testing setup, as well as in wastewater samples which provide virus and variant surveillance at a local and national level. This has the potential to boost the speed and efficacy of the public health response to new and emerging SARS-CoV-2 VOCs. Such sequence-specific tests, including nucleic acid amplification and CRISPR-Cas cleavage techniques, rely on sequence complementarity between the primers/probes and the surveyed sequence. These methods are facile, highly efficient and can rapidly identify mutations in the genomic sequence probed.

The gold-standard genetic test is qPCR and numerous SARS-CoV-2 assays have been described[2], targeting multiple mutated loci. However, the bottleneck in reagents and consumables, as well as the need for point of care testing during the pandemic has spurred the development of alternative strategies. Of particular interest has been RT-LAMP [3] which can rapidly amplify RNA at a single, constant temperature with high specificity and sensitivity. However, achieving the specificity required to discriminate single-point mutations using RT-LAMP is challenging, but the emergence of CRISPR-Cas based detection has the potential to achieve this with isothermal nucleic acid amplification in a one-pot reaction[4].

The discovery of CRISPR (Clustered Regularly Interspaced Short Palindromic Repeats) adaptive immunity in bacteria and the associated nucleases that cleave foreign RNA/DNA, has led to the development of an increasing number of highly specific genetic tests. CRISPR-Cas-based detection utilises CRISPR associated enzymes (Cas) specific to a target nucleic acid; Cas13 for RNA, and Cas12 and Cas9 for DNA or cDNA (by converting RNA using reverse transcriptase). Many of these techniques have been developed commercially for SARS-CoV-2 detection including SHERLOCK (Specific High-sensitivity Enzymatic Reporter un-LOCKing) [4–6] and DETECTR (DNA Endonuclease-Targeted CRISPR Trans Reporter) [7].

Given the threat of VOCs to vaccine efficacy, and their more general threat to at risk populations, any interventions that will help rapidly identify and limit the introduction and spread of such variants, or enable the acceleration of the public health responses, can be an important tool in public health management. Consequently, rapid variant-specific testing is a high priority challenge. Despite this, none are yet widely implemented except for the TaqPath qPCR assay, which serendipitously possesses S-gene dropout behaviour with Alpha [8] and some Omicron variants. This coincidental property of the qPCR assay with these variants has proved to be a valuable proxy to rapidly highlight likely introductions, enabling enhanced testing, contact tracing and isolation and the rapid assessment of case growth. However, this assay is only employed in a sub-section of testing labs. Whilst, the S-dropout is otherwise non-specific, its utility in early tracking of these variants clearly highlights the value of direct variant detection.

Based on the recently developed CRISPR-Cas assay SHERLOCK[5], here we develop a readily adaptable and rapid molecular diagnostic strategy applicable to all existing VOCs, that can be readily employed for newly identified variants. In combination with the power of genomic sequencing, this can aid public health management and allow timely and informed interventions that can help minimise the risk of concerning immune escape variants becoming established.

We describe the development of a generalisable approach for variant of concern molecular detection (VarLOCK). We describe optimisation for each mutation of concern, and importantly demonstrate validation and implementation with the Cardiff University COVID19 Testing Service. We demonstrate the ability to employ these assays with national wastewater surveillance to identify and map temporal and geographic population-level variant prevalence. We show this approach, when combined with LAMP amplification, could be employed widely for near-patient variant-specific diagnostic testing. The rapid adaptability of VarLOCK for new and emerging VOCs is demonstrated with the development of an Omicron specific assay within 2 weeks of WHO designating it as a VOC. This may contribute to current public health efforts by enabling more rapid identification of Omicron infection, as well as providing a generalisable approach for future VOCs.

## Results

### Concept of the VarLOCK assay

SARS-CoV-2 variants are characterised by specific sets of mutations selected in the course of virus evolution. Sub-variants diverge from the main VOC clades through acquisition of additional mutations, and as some mutations are not fully penetrant individual samples can have diverged mutational profiles. By identifying the mutations present in the sample’s genome, the variant can be identified. Mutations may be unique and associated with a single VOC, but many other mutations are shared between different VOCs potentially associated with the selective advantage they confer. In particular, variants can be differentiated by the composition of mutations in the spike protein gene (S-gene) (Table 1, Figure 1B).

**Table 1.**
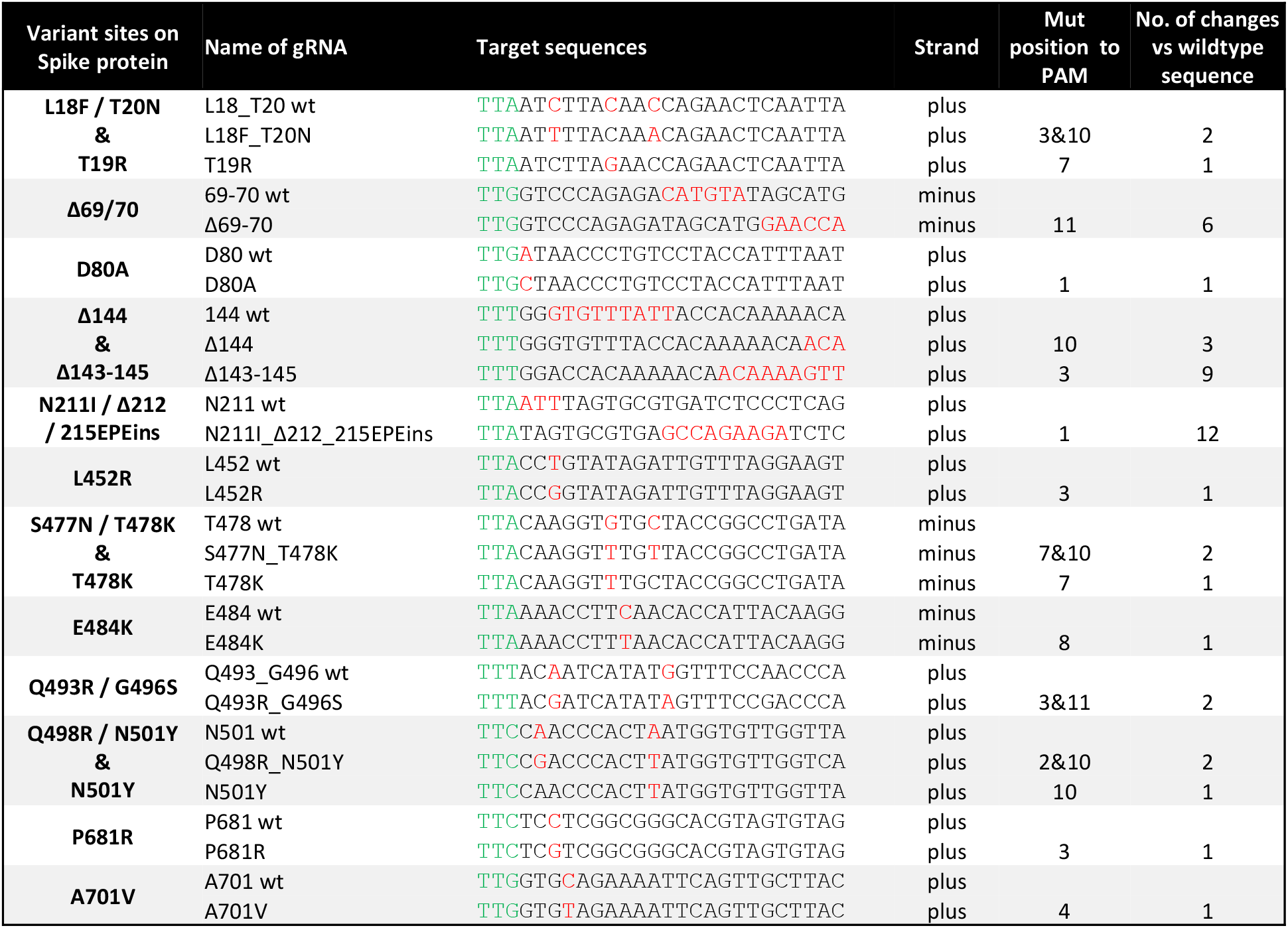
gRNA target sequences surrounding the variant sites. The PAM sequence is coloured in green. The nucleotides substituted in the variants and their corresponding nucleotides in the original Wuhan SARS-CoV-2 sequence are coloured in red. Both deleted nucleotides and the shifted sequences at Δ69-70, Δ143-145, Δ144 and Δ212 are also coloured in red.

**Figure 1.**
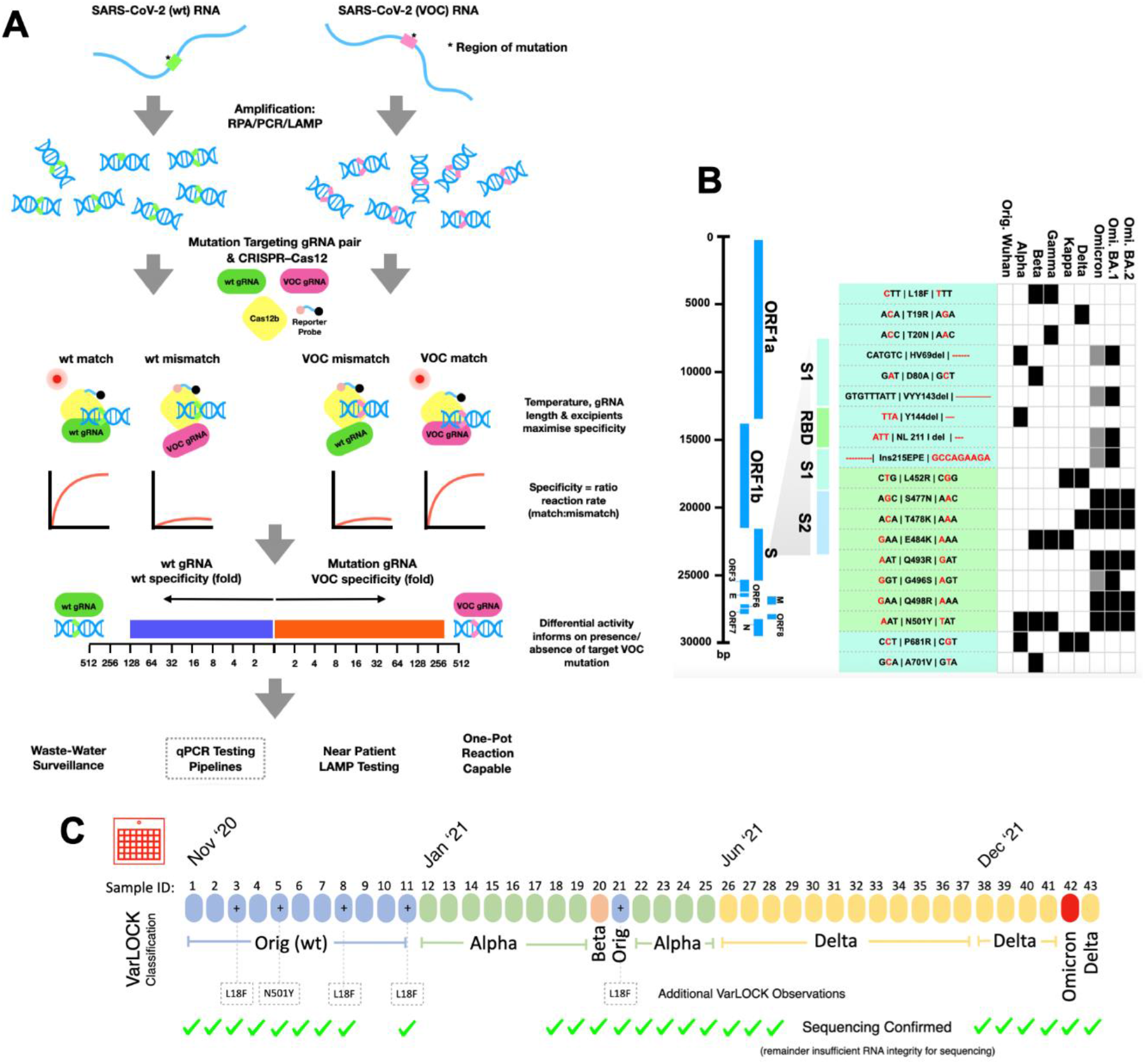
SARS-CoV-2 Variant of Concern identification by VarLOCK assay. **A)** illustration of the VarLOCK assay. The region with wildtype (green) or mutant (pink) sequence of SARS-CoV-2 RNA is amplified by either RPA, LAMP or PCR. Amplified DNA fragments are subjected to SHERLOCK assay with a pair of gRNAs matching the wildtype sequence (wt gRNA, green) or the mutant sequence (VOC gRNA, pink). We aimed to find conditions in which only a perfect match between the gRNA and the amplified DNA can trigger a collateral nuclease activation of the Cas12b protein to cleave a quenched fluorescent reporter. This allows nuclease activity to be monitored by the increase in fluorescence. The ratiometric response of an unknown SARS-CoV-2 variant sample indicates the presence, or absence, of the targeted mutation and thus enables identification of the VOC. **B)** Schematic of the genome of SARS-CoV-2 (left) with blow-up of the Spike protein encoding region showing the location of RBD, S1 and S2. Nineteen mutation sites were chosen for VarLOCK assay for which gRNA pairs were designed and optimised. Nucleotide sequences encoding the relevant amino acids are shown in the table and the nucleotide substitutions/deletions are marked in red. The association of the mutations with various variants of concern is indicated in the table, creating a barcode-like identification matrix. (C) Application of this approach to saliva samples successfully identifies original Wuhan, Alpha, Beta, Delta and Omicron variants with results confirmed by sequencing.

CRISPR cleavage is sequence specific and guided by the targeting sequence of the guide RNA (gRNA), and thus by comparing the cleavage using wildtype and mutant specific guide sequences, we reasoned that the mutational state of the investigated region could be determined. We designed primers to amplify the target sequences and gRNAs to specifically match wildtype and mutant sequences to enable the SHERLOCK method to cleave with Cas12b quenched fluorescent probes only when a specific match is present. We designed assays for various S-gene mutations namely L18F/T20N, T19R, Δ69-70, D80A, Δ143-145, Δ144, N211I/Δ212/215EPEins, L452R, S477N/T478K, T478K, E484K, Q493R/G496S, Q498R/N501Y, N501Y, P681R and A701V to enable VOCs to be detected. To account for potential non-specific cleavage and reporting, a second reaction with the wildtype sequence specific gRNA was run in parallel, with the differential response informing on the likely presence or absence of the target mutation.

### Selection of variant-defining mutation sites in the S gene

Mutations of SARS-CoV-2 are acquired during genome replication and occur randomly across the whole genome. The SARS-CoV-2 spike (S) protein plays a crucial role in receptor recognition through the receptor-binding domain (RBD), followed by membrane fusion. Mutations in the spike protein likely account for the increased transmissibility and may lead to decreased protection from immunisation and natural infection[9]. We therefore focused for VarLOCK on the differential mutations located in the S gene.

First, to identify the variant-specific Spike protein mutations in the SARS-CoV-2, we compared the S-gene sequences found in Alpha, Beta, Gamma, Delta and Omicron VOCs and Kappa (VOI - B1.617.1) and selected 19 mutation sites that are VOC characteristic and contain a Cas12b protospacer adjacent motif (PAM) site (TTN) in their vicinity to allow CRISPR-based detection assays (Figure 1A). We then designed 16 pairs of gRNAs for the 19 selected mutation sites (Table 1). Each gRNA pair targets the same region of the S gene, comprising the sequence present in either the Wuhan strain (wt) or the VOCs specific mutation (mut) (Table 1).

### SHERLOCK offers limited discrimination of single nucleotide variation

First, we tested whether the SHERLOCK assay provides sufficient discrimination for our VarLOCK approach to differentiate specific variant mutations. We performed SHERLOCK detection assays using wildtype (wt) and mutant (mut) specific gRNAs on short synthetic DNA templates containing either wildtype or mutant sequences for the tested SARS-CoV-2 genomic loci, using the reported assay conditions[5] (Supplementary Table 2 and Figure 2). For each gRNA pair, four sets of assays were performed (gRNA/template) - wt/wt and mut/mut, to check the on-target performance of the assay, as well as two discordant sets with wt/mut and mut/wt combinations to check for assay crosstalk (specificity). In an ideal case, the assays should provide high reporter activity on matching pairs with only background signal observed for discordant pairs. The detection reactions were followed in real-time by observing an increase in HEX fluorescence coming from the cleaved quenched fluorescent reporter and the reaction velocities were extracted for each of the reactions.

All the assays provided efficient on-target activity, but different targets showed varied levels of discrimination between the discordant pairs. The Δ69-70, Δ144 and P681R assays differentiated the wt and the mutant alleles effectively (>10-fold discrimination). However, other assays offered only limited discrimination (<4 fold) (i.e., D80A or T478K) or failed to discriminate wildtype and mutant templates (i.e., E484K, L452R or N501Y). Of the 8 pairs of gRNAs for the variants containing single nucleotide substitution, only two (T19R and P681R) showed clear discriminative activities between matched and mismatched targets (Figure 2). Not surprisingly the position of the substitution on the gRNA target sequences relative to PAM and the number of nucleotide exchanges affected the degree of discrimination as observed for the other CRISPR/Cas systems (Table 1). The two deletion sites, Δ69-70 and Δ144, have the greatest power to discriminate the mut from wt. Single nucleotide exchanges could only be efficiently discriminated in the P681R assay, for which the substitution is only 3 bp away from PAM. Interestingly, D80A, for which the substitution is right next to PAM did not show strong discrimination between the wildtype and mutant templates.

**Figure 2.**
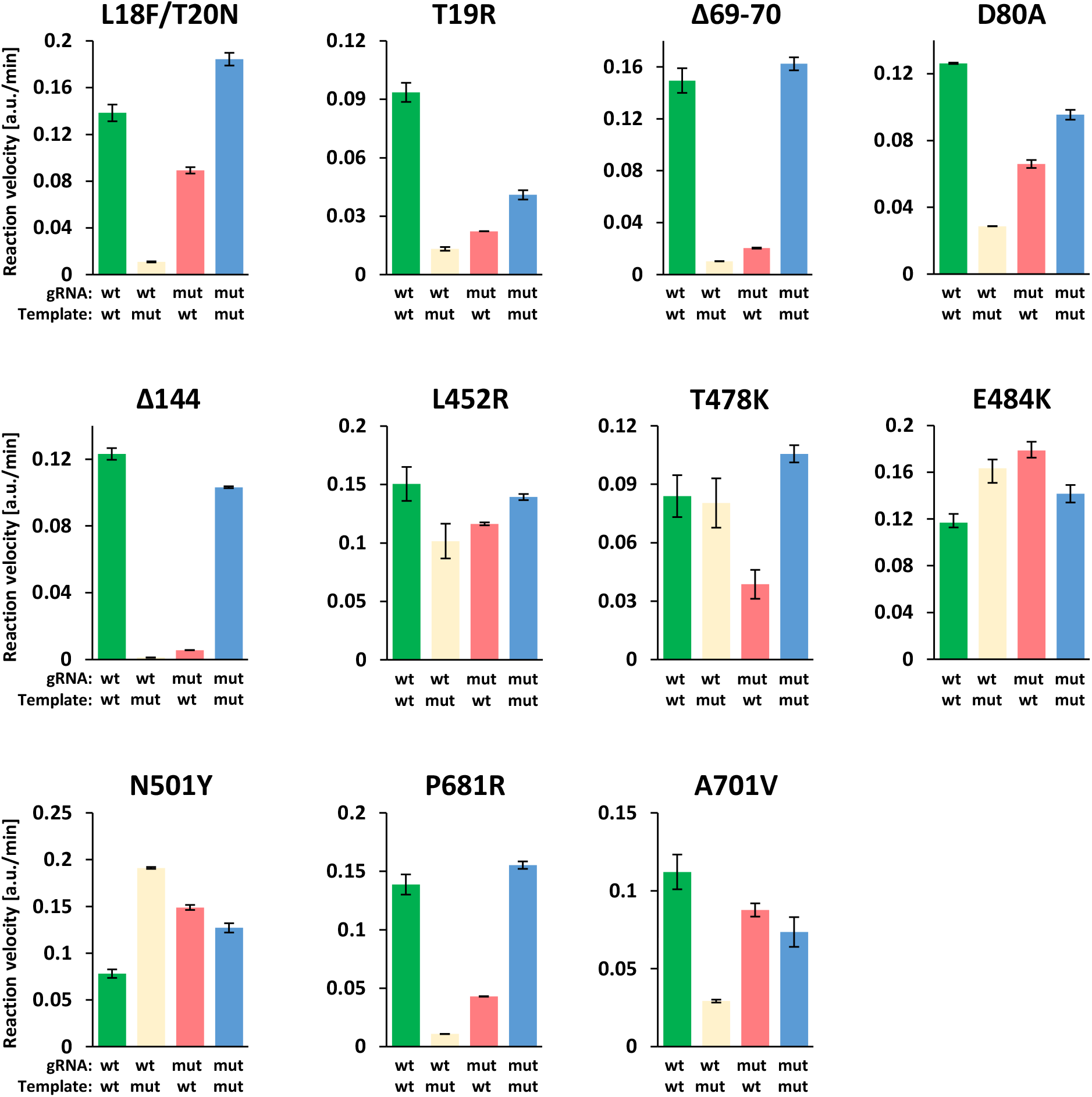
Detection specificity is low for single nucleotide substitution with standard conditions. A representative experiment showing collateral nuclease activities (y axis; arbitrary fluorescence unit increase/min) triggered by wt gRNA matching wt target (wild type detection), wt gRNA matching mutation target (wt cross-reaction), mutation gRNA matching wt target (mutation cross-reaction) and mutation gRNA matching mutation target (mutation detection). Mutations are indicated on the top of each panel. Average and SD are derived from technical duplicates.

### Assay optimisation for efficient discrimination of variants

As observed, the “standard” SHERLOCK conditions afforded only limited discrimination of the VOC-specific mutations. Whereas more extensive genetic changes could be discriminated, single nucleotide exchanges distal to the PAM could not. We therefore surveyed various assay conditions and chemical additives to increase the discriminatory power of the assay. The base-pairing specificity of nucleic acids can be influenced by numerous factors. For example, in PCR reactions, the specificity of primer binding to the template is, amongst other factors, dependent on the annealing temperature used and the reaction ionic strength. We have limited the search to conditions that can be tolerated by the polymerases and enzymes used in the assay to permit a “single pot” reaction.

We selected several common PCR enhancers that are used to alleviate specificity issues in PCR amplification: DMSO, trehalose, Gly-Gly, betaine, 1,2-propanediol, as well as GITC which have been shown to improve amplification efficiency and/or specificity[10–12]. Furthermore, we checked whether specificity could be enhanced by increasing assay temperature or by using gRNAs with a shorter spacer sequence as previously observed for CRISPR/Cas9 specificity[13].

### PCR enhancers show limited assay selectivity improvement

We selected the gRNAs pairs for L18F/T20N (g18/20 wt and g18/20 mut) and the E484K (g484 wt and g484 mut) as examples for testing various conditions (Supplementary Figure S1 and S2C). The PCR enhancers including: DMSO, Gly-Gly, betaine (up to 1 M), trehalose, 1,2-propanediol, or a combination of these, did not appear to improve significantly VarLOCK specificity (Supplementary Figure S1A-B), neither did the NEB Q5 high GC enhancer.

Strikingly, addition of salts at increasing concentrations improved template discrimination. Increasing salt concentration in the reaction mixture of KCl to 150 mM or supplementing with 40-50 mM GITC significantly improved the specificity for E484K (Figure 3A and Supplementary Figure S1B-C). KCl and GITC at a similar concentration also improved N501Y specificity (Figure 3A and Supplementary Figure S1D). Surprisingly, both KCl and GITC at similar concentration did not improve the specificity for A701V, but inhibited Cas12b activity altogether (Figure 3). Taurine at 200 to 250 mM concentration improved specificities moderately for L18F/T20N and E484K (Figure 3A), but no beneficial effect was observed for A701V (Figure 3A). Overall, chemical additives only moderately improved VarLOCK specificities for some variant sites but did not show a general effect.

**Figure 3.**
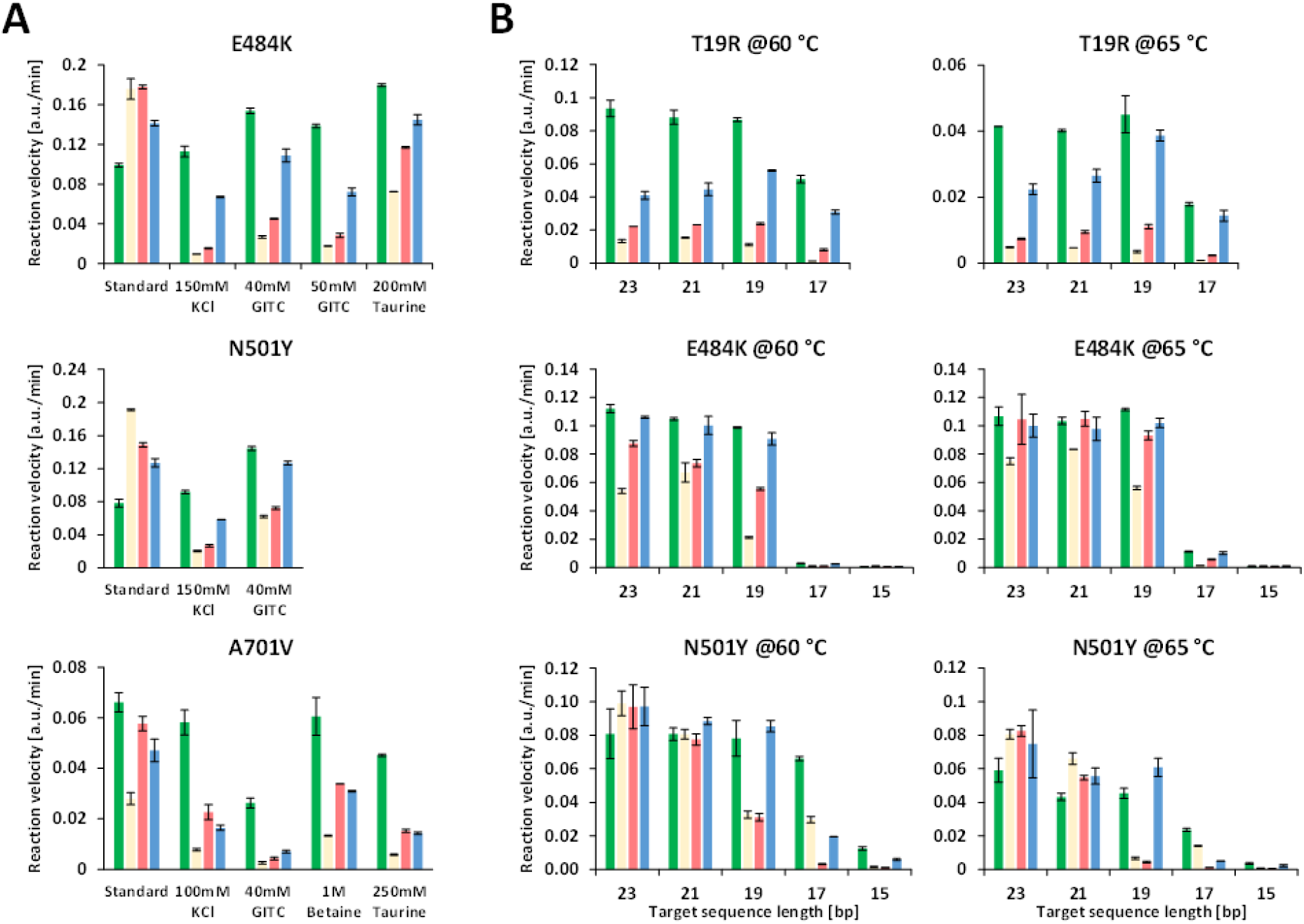
Detection specificity can be improved by interfering stability of intermolecular interaction, gRNA length and reaction temperature. **A)** Reactions were performed in standard buffer or with the indicated concentrations of additives. Reporter activities triggered by wt or mut gRNA matching or mismatching the targets are calculated as increase of arbitrary fluorescent unit/min. Representative experiments for mutation E484K, N501Y and A701V are shown. Average and SD are derived from technical duplicates. **B**) reaction performed at 60°C or 65°C. The lengths of target sequences in gRNA are indicated under the x axis. Average and SD are derived from technical duplicates of a representative experiment.

The effect of these salts suggests that the low ionic strength of the buffer may contribute to the lack of specificity, or that unspecific charge interactions between DNA and the ribonucleoprotein (RNP) (Cas12/gRNA) promote stability of the mis-coupled substrate/RNP complex.

### Shorter gRNAs and increased assay temperature improve selectivity

Decreasing the length of the gRNA spacer sequence has previously been shown to improve the specificity of Cas9 cleavage [13]. We tested whether a similar effect for Cas12b could be observed and that shortening the length of the gRNA spacer sequences might increase relative base-paring specificities through reduction of free energies of annealing between gRNA and its target.

We tested assay activity and specificity for 7 variant sites with spacer lengths between 23 and 15 nucleotides in reactions performed at two different temperatures. At 60°C, activities remained comparable for spacer lengths between 19- and 23-nt for all 7 variant sites tested (Figure 3B and Supplementary Figure S2A, C, E, G). Specificities were increased with 19-nt spacers in gRNA for T478K, E484K and N501Y, without affecting the on-target signal strength. Further shortening of the spacers to 17-nt dramatically decreased the activities, except those for L18F/T20N, T19R, L452R and P681R. None of the gRNAs with a spacer length of only 15-nt supported cleavage activity (Figure 3B and Supplementary Figure S2A, C, E, G).

Alternatively, increasing temperature could reduce annealing stabilities of mismatched counterparts between gRNA and targets, with minimum effects on matched counterparts. As AapCas12b can tolerate temperatures as high as 100°C [14], we tried to rerun the reactions using the gRNAs with difference length of spacers at 65°C (Figure 3B and Supplementary Figure S2B, D, F, H). In comparison with the reactions at 60°C, increasing assay temperature by 5°C did improve specificities significantly. The most improved variant site was N501Y detected using gRNAs with a 19-nt spacer (Figure 3B).

### Combination of approaches

As high specificity detection for variant sites L452R and T478K were not achieved by either altering gRNA length or temperature, we decided to test if KCL, GITC and taurine at concentrations that were optimal for E484K and N501Y could also provide a benefit in combination with gRNA length (Supplementary Figure S3A and B). For L452R gRNAs with 21-, 19- and 17-nt spacers, both GITC and KCl drastically improved specificities with moderate reduction of activities as a compensation (Supplementary Figure S3A). Taurine also increased specificities, but to a lesser extent compared to GITC and KCl, however. We calculated the ratio of activities derived from gRNA/Target-matched and mismatched reactions as an indicator of specificities (Supplementary Figure S3B). We saw that the effects of the chemicals were both on gRNAs that match the wildtype sequence (g452wt) and the variant sequence (g452mut). To evaluate the overall specificities, we took the averages of the two ratios (g452wt and g452mut) as a combined fold difference for comparison across all reaction conditions. Almost 8-fold discrimination was achieved for 21-nt gRNA with 50 mM GITC and 21-nt and 17-nt gRNA with 150 mM KCl (Supplementary Figure S3B). Similarly, testing on T478K showed that all the three chemicals improved specificities (Supplementary Figure S3C). In this case, activities remained at similar levels in reactions with GITC, taurine and KCl, compared to the standard reaction, at least for 23- and 21-nt gRNAs (Supplementary Figure S3C). GITC and KCl reduced the activities slightly for 19-nt gRNAs. The overall fold-differences for matched/mismatched reaction reached 10-fold with 21-nt gRNA with both GITC and KCl (Supplementary Figure S3D).

Taken together, for the variant sites with more than two nucleotide changes, L18F/T20N, Δ69-70 and Δ144, specificities can be achieved in standard buffer at 65°C with 23-nt gRNA (see a summary in Figure 4). Two sites with only single nucleotide substitutions, T19R and P681, can also work well in standard reaction run at 65°C with 23-, 21- and 19-nt gRNAs targeting sequences. L452R, T478K and N501Y require shorter gRNA, chemical additives and higher temperature to achieve specificity with overall fold-difference above 7.

**Figure 4.**
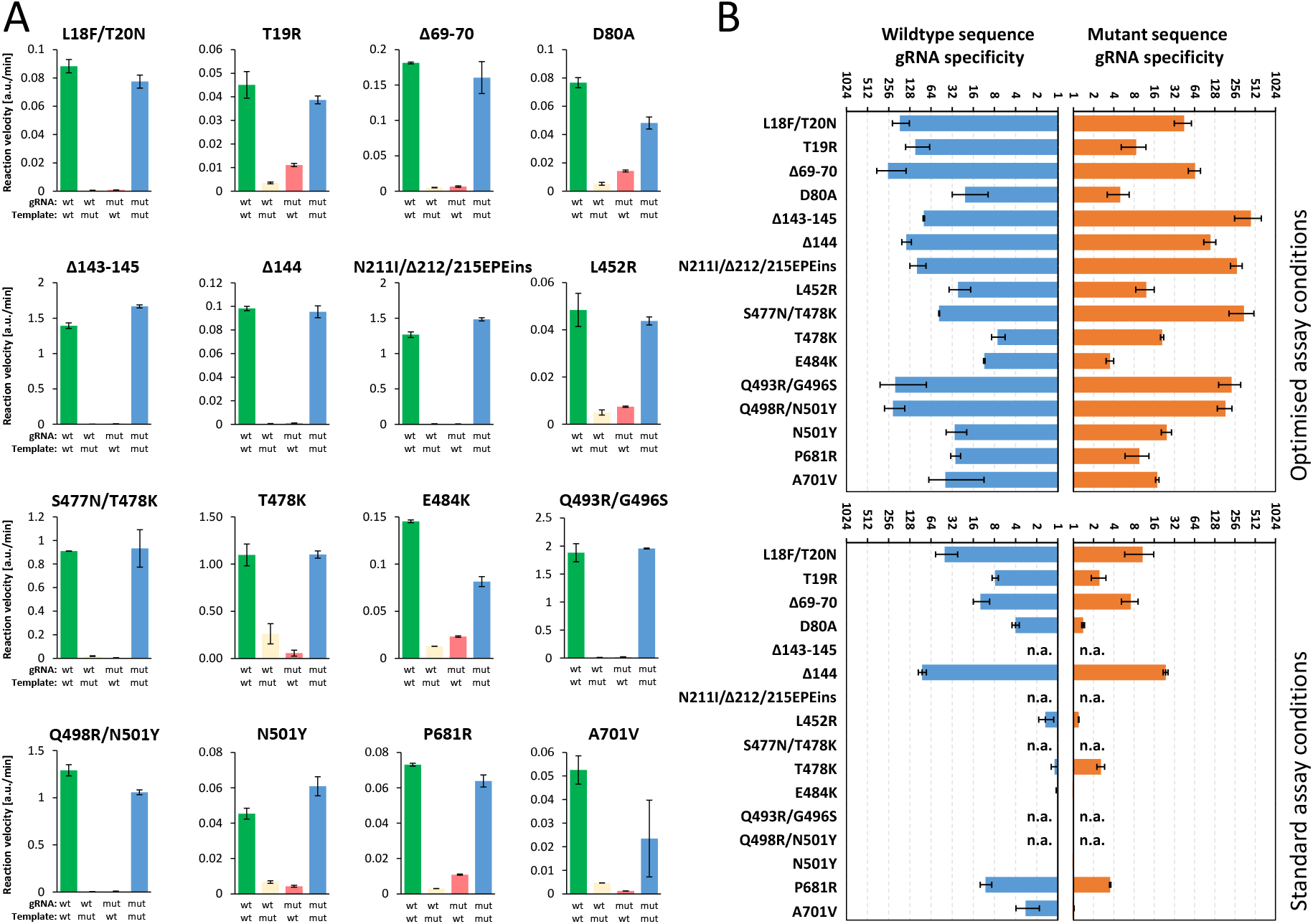
VarLOCK assay sensitively discriminates single nucleotide exchanges. **A)** Specificity plots for representative examples for each mutation performed under optimised conditions (as listed in Table 2). Average and SD are derived from technical duplicates. **B)** Specificity factors of the gRNA (wt blue bars and mutation in orange) are presented as ratio of the two reaction activities triggered by matched and mismatched target DNA. Please note that the ratios are displayed on a log scale. The bar graph shows average, and SD of the ratios derived from 2-5 biological repeats.

### Adaptation of the VarLOCK with lateral flow assay

After optimisation of the VarLOCK assays with fluorescence detection, which can be easily applied to large scale screening, we asked whether this assay could be adapted to point of care lateral flow assay. We chose two mutation sites, Δ69-70, which probes for a 6-nucleotide deletion, and N501Y with just a single nucleotide substitution. As shown, specific detection had been clearly observed for both assays. A very faint non-specific band developed in both no target control (N) and mismatch targets, which may reflect the property of the batch of the dipstick used. Quantitative image analysis of band intensity found equivalent signal in these two scenarios in comparison to the VOC match (Supplementary Figure S5).

### Validation of the VarLOCK assay using saliva samples

Next, we sought to check the performance of the VarLOCK assay on positive saliva samples identified at Cardiff University’s COVID-19 screening service. We randomly selected 11, 14 and 15 positive samples collected in Nov 2020, between Feb and May 2021, and between Jun and Aug 2021, respectively. Based on the historical data, in the three selected time periods, the Wuhan, Alpha and Delta variants were dominant in the population, respectively. We used the VarLOCK assay to probe the presence of 6 mutations that should provide sufficient information to identify the variant present. Among these, we selected one mutation specific for each variant: Δ144 for Alpha, A701V for Beta, L18F/T20N for Gamma and T19R for Delta variants. We also included two additional mutations found within the RBD region of the S gene, T478K, which is unique for the Delta variant, and N501Y, which is shared between Alpha, Beta and Gamma variants. These mutations are suggested to be responsible for increased infection rates and reduced protection by vaccination and antibody therapies[15, 16].

The relevant regions of interests were amplified by RT-PCR from saliva samples and the products were subjected to VarLOCK assays. The results are presented as Log10 of the ratio of the two VarLOCK activities obtained with wt gRNA and mut gRNA. This provides a simple method to visualise samples as wildtype or mutant, based on whether the Log10 value is above or below 0 respectively (Figure 5). This presentation was evaluated with wildtype and mutant control targets, and both 1 and 10 nM control targets showed similar Log10 value, indicating a tolerance of variabilities of RT-qPCR amplification in all 6 mutation sites (Figure 5).

**Figure 5.**
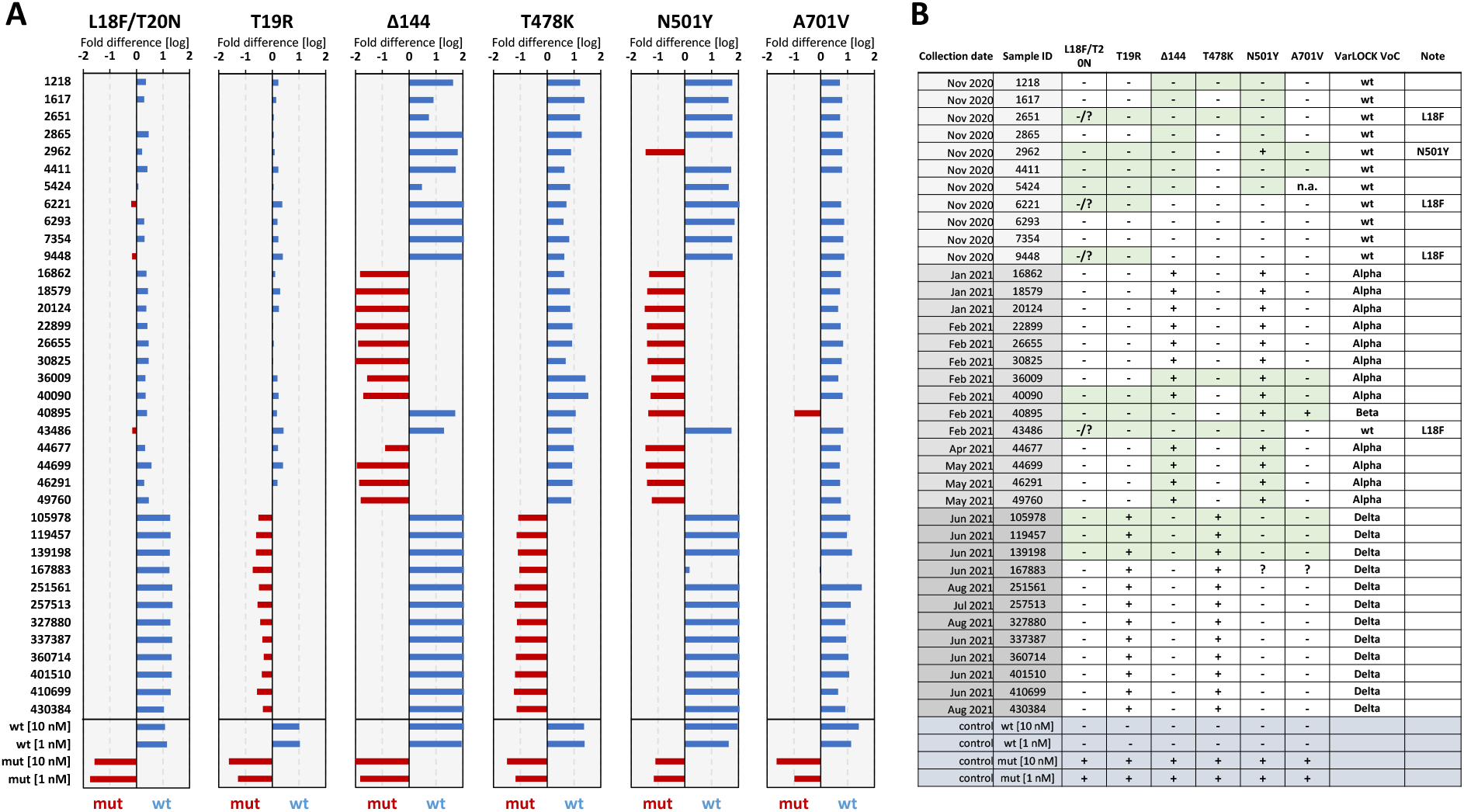
Variant identification of the positive saliva samples. **A)** VarLOCK assay results showing dominant wildtype signal (blue) or mutant signal (red), calculated as Log_10_ of fold (wt/mut) differences. Samples are grouped in three time periods as indicated with different shading of greyness. **B)** VarLOCK detection summary for the analysed samples with VOC assignment. Sanger sequencing was used to confirm the variants for samples and amplicons shaded in green.

Δ144, a signature for Alpha variant, showed that 12 out of 14 samples collected between February and May 2021 were assigned as Alpha. These 12 samples also carried the N501Y mutation as typically found in Alpha (Figure 5). The presence of A701V mutation (Beta) was only detected in one sample (40895). This sample also carried the N501Y mutation. The A701V assay also produced ambiguous results for 167883. To understand the cause of the ambiguity, we sequenced this sample. Sequencing suggested that, although this sample had a small portion of C to T substitution, the sample may contain more than one sequences (Supplementary Figure S7).

The L18F/T20N assay detecting a Gamma specific mutation, did not indicate the presence of this mutation in any of the samples tested, nor did it conclusively assign the wt sequence in 4 of the tested samples, namely 2651, 6221, 9448 and 43486, suggesting the presence of a different mutation from L18F/T20N. Indeed, Sanger sequencing of these PCR products showed that all these 4 samples carried the spike protein L18F mutation (single nucleotide substitution from C to T) (a representative sequencing result shown in Supplementary Figure S7B). The Delta variant probe T19R showed that all 12 samples had this mutation (Supplementary Figure S5). Sanger sequencing of a few selected templates was used to validate the VarLOCK results (Supplementary Figure S6). Indeed, samples which tested positive for mutation in the VarLOCK assay contained the expected sequences when sequenced (Figure 5B). Interestingly, although no VOCs were designated before December 2020, we detected 3 samples with the L18F mutation out of the 11 samples collected in November 2020. Twelve of the 14 samples collected between January and May 2021 were Alpha variants and all the 12 samples collected between June and August 2021 were Delta variants.

### VarLOCK wastewater

Wastewater monitoring for the detection of SARS-CoV-2 has become an important tool, complementing hospital and community-based clinical surveillance to identify the presence and relative rates of transmission of COVID-19 infection within the population of a defined geographical area [17]. Wales was one of the first countries to implement nation-wide surveillance of COVID-19 in wastewater, which now covers all of Wales’ major urban areas and ~80% of the Welsh population[18]. Globally, there is increasing interest in local and national wastewater surveillance to monitor the effect of public health interventions and monitor or inform of possible new local outbreaks ahead of, or as an alternative to, mass testing. We hypothesised that the VarLOCK assay could be used to monitor and map the prevalence of variants of concern in close to real time. In principle, this could provide speed, coverage or resource advantages over symptomatic testing and subsequent sequencing of selected positive cases. This could be especially useful where sequencing resources are scarce or when national COVID-19 caseload is high in a highly vaccinated population, but the emergence or introduction of immune-escape variants with known mutations of concern could risk an increase in severe disease. To evaluate the feasibility of applying VarLOCK to identify VOCs in wastewater, VarLOCK assays were used to discriminate between the original Wuhan, Alpha, Beta and Delta variants retrospectively in wastewater samples collected from three wastewater treatment works (WwTWs) serving the cities of Cardiff, Swansea and Newport (all urban centres in South Wales), which were collected weekly from November 2020 to November 2021, as part of the COVID-19 Welsh Wastewater monitoring programme.

Unlike samples isolated from an individual, where the presence of only one SARS-CoV-2 variant is normally expected, wastewater samples contain a mixture of numerous individual samples. In addition, in contrast to samples isolated from an individual, the viral genomes in the wastewater are highly fragmented, and the samples are complex and contain nucleic acids from a wide range of sources, posing a hurdle for sequencing. Weekly samples were processed with the VarLOCK assay to determine the prevalence of different variants. For this the relevant regions were amplified with RT-PCR and the products analysed with assays probing for Δ69-70, Δ144 (Alpha), T478K (Delta) and A701V (Beta).

For the Cardiff, Newport and Swansea samples, a differential VarLOCK mutation-specific signal is observed with time, reflecting the respective introduction and then dominance of the original variant, Alpha and then Delta VOCs in the country (Figure 6A). The sequential introduction and growth of the Alpha variant in Cardiff, followed by Newport and then Swansea is notable and likely reflective of the tightening of Government COVID19 restrictions in Wales through December 2020 and January 2021, with limited household mixing, stay-at-home orders and restricted travel accounting for a slow-down in seeding events with reduced inter-city mixing. In contrast Delta VOC arrival and dominance appears more uniformly across the cities with this variant becoming established in the U.K. amidst lighter restrictions. Across the three cities the combined VarLOCK VOC profile maps extremely well to the all-Wales genomic surveillance data catalogued by the COVID-19 Genomics UK Consortium (COG) (Figure 6B top panel). Positive SARS-CoV-2 detection and VOC identification is also made when community prevalence is low (e.g. June 2021) indicating the sensitivity of the approach.

**Figure 6.**
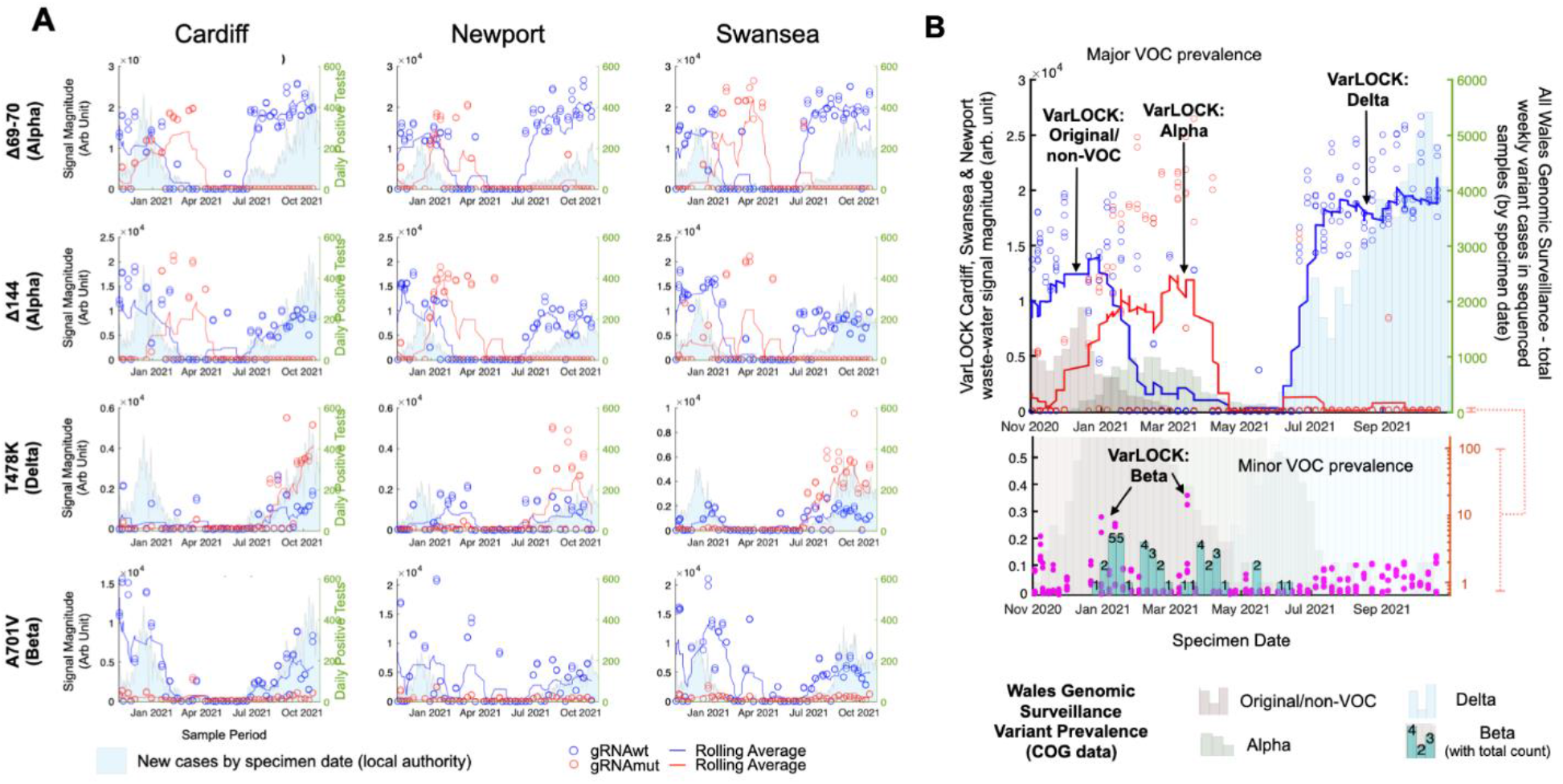
VarLOCK analysis of wastewater samples. Wastewater samples collected weekly between November 2020 and November 2021 at three urban centres in south Wales were analysed with VarLOCK assays for original variant, Alpha, Beta and Delta VOCs. The results showed successful application of VarLOCK for variant monitoring in wastewater with the VarLOCK variant signal reflecting dominant periods of the original variant, Alpha and then Delta VOCs in the country (panel B). The growth of the Alpha variant can be seen to arrive first in Cardiff, followed by Newport and then Swansea (panel A). Against a high background of Alpha variant VarLOCK signal indicative of the presence of the low prevalence Beta variant is observed in the wastewater samples correlating with sequencing identified clusters in Wales (panel B).

**Figure 7.**
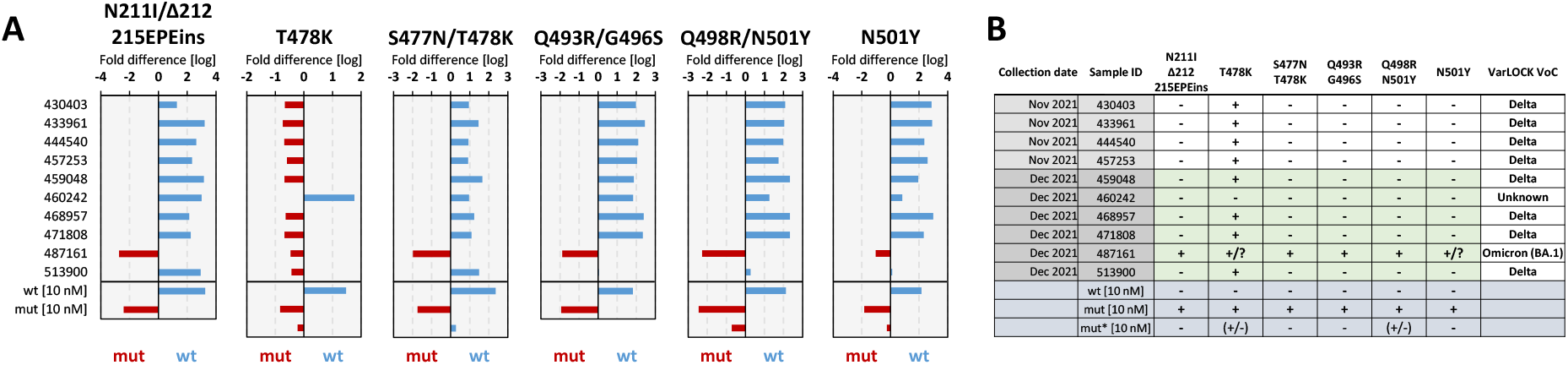
VarLOCK identification of Omicron variant in saliva samples. **A)** VarLOCK assay results showing dominant wildtype signal (blue) or mutant signal (red), calculated as Log_10_ of fold (wt/mut) differences. For the regions where mutations are present in other VOCs, an additional control was used with alternative template for the other mutants (mut*) **B)** VarLOCK detection summary for the analysed samples with VOC assignment. Illumina sequencing was used to confirm the variants for samples shaded in green.

Precise caseload quantification by waste-water monitoring is known to be challenging[17]. For example, here some wastewater samples recorded no observable viral material despite being collected during a period of known high prevalence. We attributed this to variability of conditions and/or interplay of timing in the wastewater-processing and sample collection and this would require further investigation (these null data points have been retained in the presented data to avoid subjective bias). However, VarLOCK VOC identification in wastewater is observed to be effective at both high and low community caseload throughout the study. Further to this, we observed VarLOCK identification of the presence of mutations characteristic of the Beta variant in wastewater samples. Community transmission of Beta VOC was limited in Wales with the variant not becoming established in the presence of high Alpha caseload and restrictions reducing transmission, with only 40 cases across all of Wales recorded by sequencing during this study period. The VarLOCK Beta VOC response temporally correlates with recorded sequence clusters in Wales (Figure 6B lower panel). Here, the VarLOCK VOC assay was able to identify the presence of the sparse Beta variant against a backdrop of high Alpha caseload, showing the sensitivity of the approach. Whilst this study was retrospective, collectively these data highlight the promising potential for utilising wastewater VarLOCK monitoring for near-real-time VOC monitoring.

### Detection of Omicron variant and subvariants

Recently, a novel SARS-CoV-2 variant B.1.1.529 was identified in Southern Africa and flagged as containing a potentially concerning number of spike mutations on 23^rd^ November 2021 following sequence identification in Botswana (3 genomes), Hong Kong ex S. Africa (1 genome, partial) [19]. South African public health recorded an extremely large and rapid increase in SARS-CoV-2 cases and test positivity in Gauteng Province for the week of 14-20 November 2021 [20] with this rapidly attributed to Omicron by South Africa Centre for Epidemic Response & Innovation [21, 22]. It was designated by WHO and named Omicron VOC on 26^th^ November 2021. Local epidemiology studies showed significant re-infection risk in a population with relatively low vaccination coverage [23] and early laboratory studies indicated significant reduction in neutralising ability of vaccine and prior-infection induced antibodies [24–26]. This reinfection and incomplete vaccine protection risk has been borne out in populations of higher vaccination coverage following Omicron introduction in the U.K. with the variant rapidly becoming dominant against a background of high Delta caseload. The rapid spread of Omicron across the globe highlights the global public health challenge in containing VOCs following their identification. Whilst sequencing has been indispensable in the scientific effort in monitoring, tracking, understanding and tackling Omicron, the associated lag has hindered containment or implementation of targeted proactive measures, where rapid molecular diagnostics could arguably provide important speed and scale advantages. S-gene target failure (SGTF) has proved a useful proxy for this, enabling rapid assessment of early Omicron case growth in U.K. (2-3 day doubling) [27] and determining dominance in England by 14^th^ December [28, 29]. However, SGTF is currently limited to four main qPCR laboratories in the U.K. (Alderley Park, Glasgow, Milton Keynes, and Newcastle Lighthouse Laboratories) for community (Pillar 2) testing, so many areas and cases remain under-reported until community transmission is well established. Furthermore STGF occurs due to fortuitous primer design, and cannot be relied upon for identification of future variants.

In this context we applied our generalisable VOC detection strategy to Omicron, developing and implementing an assay able to differentiate Omicron from delta, wt and other variants and additionally positively detect and distinguish between Omicron sub-variants BA.1 and BA.2, where BA.2 is undetectable by SGTF. Within 4 days from receiving the oligonucleotides, we successfully developed 5 VarLOCK assays to probe for the presence of Omicron specific mutations in the S gene: Δ143-145 (BA.1), N211I/Δ212/215EPEins (BA.1), S477N / T478K (BA.1 and BA.2), Q493A/G496S (BA.1) and Q498R/N501Y (BA.1 and BA.2) (Table 1 and 2). First, the assays were tested and optimized with synthetic dsDNA templates (Figure 4). In addition, given that the same S gene locations are found differently mutated in other variants, we cross-validated the specificity of the Omicron assays against the templates containing other VOC specific sequences (Figure 4). All the assays showed excellent specificity, ranging between 49.1 and 347.9-fold discrimination. Subsequently, we applied the VarLOCK Omicron detection to 10 recently isolated positive saliva samples collected between the last week of November and mid December 2021. In one of the samples - 487161, the VarLOCK assays identified the presence of N211I/Δ212/215EPEins, S477N/T478K, Q493A/G496S, Q498R/N501Y Omicron specific mutations with good confidence. The 6 most recent positive samples were selected for whole-genome SARS-CoV-2 sequencing using the NimaGen protocol. In the VarLOCK-identified Omicron saliva sample, deep sequencing reconfirmed the presence of the detected mutations and Nextstrain server assigned the sample as Omicron (21K). The other 5 sequenced samples were assigned to the Delta clade (21J), thus reconfirming VarLOCK VOC detection results obtained.

**Table 2.** A summary of detection specificities in optimised conditions. The table shows positions and amino acid substitutions in the spike protein and the corresponding nucleotide mutations (in brackets). The VOC, the gRNA sets, the target lengths in gRNA (the preferred length are shown in Bold). Reaction temperature (in Celsius) and additives (preferred additives in Bold) are indicated in the table.

| Mutations (N) | Variants detected | gRNA set | target length | Reaction temperature | Reaction conditions (additives) |
| --- | --- | --- | --- | --- | --- |
| L18F(C>T) / T20N(C>A) | Gamma | L18_T20 wt :: L18F_T20N | 23,**21**,19 | 65 °C | standard |
| T19R(C>G) | Delta | T19 wt :: T19R | 23,21,**19** | 65 °C | standard |
| Δ69-70(CATGTC) | Alpha | 69-70 wt :: Δ69-70 | 23 | 65 °C | standard |
| D80A(A>C) | Beta | D80 wt :: D80A | 23 | 65 °C | standard |
| Δ143-145(GTGTTTATT) | Omicron | 144 wt :: Δ143-145 | 23 | 65 °C | standard |
| Δ144(TTA) | Alpha | 144 wt :: Δ144 | 23 | 65 °C | standard |
| N211I_Δ212_215EPEins | Omicron | N211 wt :: N211I_Δ212_215EPEins | 23 | 65 °C | standard |
| L452R(T>G) | Delta | L452 wt :: L452R | **21**,19 | 65 °C | **50mM GITC**, 150mM KCl |
| S477N(G>A) /T478K(C>A) | Omicron | T478 wt :: S477-8mut | 21 | 65 °C | **150mM KCl** |
| T478K(C>A) | Delta | T478 wt :: T478K | **21**,19 | 65 °C | **150mM KCl**, 50mM GITC |
| E484K(G>A) | Beta<br>Gamma | E484 wt :: E484K | 23 | 60 °C | **150mM KCl**, 50mM GITC |
| Q493R/G496S | Omicron | Q493_G496 wt :: Q493R_G496S | 21 | 65 °C | standard |
| Q498R(G>A)/N501Y(A>T) | Omicron | N501 wt :: Q498R_N501Y | 23 | 65 °C | standard |
| N501Y(A>T) | Alpha<br>Beta<br>Gamma | N501 wt :: N501Y | 19 | 65 °C | standard |
| P681R(C>G) | Delta | P681 wt :: P681R | **23**,21,19 | 65 °C | standard |
| A701V(C>T) | Beta | A701 wt :: A701V | 23 | 65 °C | standard |

## Conclusions

As the COVID-19 pandemic continues, SARS-CoV-2 will continue to mutate, and novel variants will arise. It is likely further variants with transmission advantages afforded by immune escape will emerge. Once mutational sequences are known, VarLOCK provides a readily implementable means to monitor their introduction and progression. This can provide new tools in the management of the pandemic. For example, variant specific testing for travel or events; or enabling more rapid implementation of enhanced public health measures, such as surge testing or enhanced contact tracing upon early knowledge of importation or community transmission of VOCs. In this context, whilst genomic sequencing affords unparalleled insight into viral identification and lineage tracking, especially in the evolution of novel variants, it requires relatively high viral loads and is subject to delay and capacity limits, even in well-resourced nations. This lag may contribute to variants gaining an unstoppable foothold diminishing the effectiveness of public health containment efforts, as seen in the U.K. with Delta and Omicron VOCs where spread has been rapid. Consequently, faster as well as more time and resource efficient variant identification methods can contribute to more rapid and targeted public health responses when necessary.

In this regard, the development of new variant specific VarLOCK assays is rapid. The Omicron detection assays were developed within four days from the receipt of the required oligonucleotides, which can be extremely rapidly designed following identification of concerning genomic sequences. This may also be considered on the basis of predicted mutations that may be of concern to monitor for their emergence. For example, S477N, Q498R, N501Y mutations, as found in Omicron, were predicted to substantially increase ACE2 binding if to arise in combination [30]. Here, the reported Omicron VarLOCK assay was designed and tested with synthetic samples before a positive Omicron case had reached our 3000 sample/day capacity SARS-CoV-2 testing pipeline.

We show here that VarLOCK is applicable to a range of analytical samples, not only enabling variant identification in saliva samples from positive individuals as part of a PCR testing workflow, but that it is also applicable to the monitoring of variant prevalence in pooled population samples. Further, the possibility to use the VarLOCK approach with different amplification approaches (RPA, LAMP and PCR) with fluorescent or colourimetric readout means it can be integrated into PCR testing pipelines, coupled with near patient LAMP testing and is even compatible with lateral flow readout.

We have demonstrated that the developed VarLOCK approach is scalable, easily adaptable for different mutations and readily implemented on different sample types including highly fragmented pooled population samples. The VarLOCK approach is able to discriminate single nucleotide exchanges in the identification of VOCs. To achieve this level of specificity optimisation of assay conditions considering gRNA length, reaction temperature and excipients favouring binding discrimination are used to enable enhanced SNP identification. This approach significantly improves the selectivity of previously reported SHERLOCK assays, and by developing targeted VarLOCK assays with a combination of mutation specific gRNAs we are able to identify the mutational pattern present and enable successful VOC identification in infected individuals and in wastewater samples from urban populations. In this regard even low prevalence Beta VOC signatures are observed in South Wales wastewater during early 2021 amidst a background of very high prevalence Alpha infections, highlighting the detection sensitivity of the approach to new variant introductions.

The optimised VarLOCK assay is able to discriminate single nucleotide polymorphism in both DNA and RNA species. Here, we apply it to identify SARS-CoV-2 variants. We show the approach can be rapidly repurposed for new VOCs with the development and implementation of Omicron specific tests, including differentiation of BA.1 and BA.2 sub-types. VarLOCK may also be employed for sensitive and specific identification of other pathogens, for example influenza (A/B), RSV or identification of antibiotic resistance bacteria, wherever genetic identification, or especially SNP or variant discrimination, is of importance. This may contribute to future pandemic preparedness, especially as the approach is readily integrated with existing testing infrastructure. The ability to apply to population-level wastewater samples may also afford the opportunity for multi-pathogen monitoring in communities for enhanced public health surveillance.

In summary, the VarLOCK approach allows rapid implementation and development in response to new pathogens or genetic variants, with the assay being suitable for implementation at point-of-care, for integration into qPCR testing pipelines and application with wastewater surveillance. When combined with pathogen genomic sequencing VarLOCK approaches can rapidly translate sequenced variant information into rapid, sensitive, and specific nucleic acid diagnostics for enhanced public health benefit.

## Materials and methods

### Purification of AapCas12b

pAG001 His6-TwinStrep-SUMO-AapCas12b was a gift from Omar Abudayyeh and Jonathan Gootenberg (Addgene plasmid # 153162) [4]. The purification procedure of AapCas12b was adapted from the protocol described for TwinStrep−SUMO−LwaCas13a with modifications [31]. Briefly, protein expression was induced by adding 0.5 mM IPTG to transformed BL21-CodonPlus (DE3)-RIL cells (Agilent) and incubated at 18°C for overnight. The cell pellet from 1 L culture was resuspended in 40 ml Lysis Buffer (20 mM Tris pH7.5, 0.5 M NaCl, 10% Glycerol, 1 mM DTT, and 1 mM PMSF) and sonicated for 10 min at a temperature below 7 °C. Cleared supernatant was incubated with 1 ml Ni-NTA Agarose (Genaxxon Bioscience, S5377) for 1 hr at 4 °C. After incubation, the slurry was loaded on a gravity flow purification column. The column was washed with 100 ml of Wash Buffer I (20 mM Tris-HCl pH 7.5, 0.5 M NaCl and 10 mM imidazole) followed by 100 ml Wash Buffer II (20 mM Tris-HCl pH 7.5, 0.5 M NaCl and 20 mM Imidazole). The His6-TwinStrep-SUMO-AapCas12b protein was cleaved by His6-Ulp1 (in-house) in SUMO Cleavage Buffer (20 mM Tris pH 7.5, 0.5 M NaCl, 1 mM DTT and 0.15% Igepal CA-630) at 4°C overnight. Cleaved AapCas12b was eluted in SUMO Cleavage Buffer and dialysed against Dialysis Buffer I (10 mM Tris-HCl pH 7.5, 0.2 M NaCl, 1 mM DTT and 10% glycerol) for 2 hr and Dialysis Buffer II (10 mM Tris-HCl pH 7.5, 0.1 M NaCl, 1 mM DTT and 50% glycerol) for 3 hr. Protein purity and concentration were estimated with SDS-PAGE.

### gRNA design, synthesis and purification

As the AacCas12b scaffold-based sgRNA has been reported to produce more robust and specific nuclease activity in combination with AapCas12b compared to previously used gRNAs, we designed our gRNA with the AacCas12b gRNA scaffold [4]. In order to synthesise gRNA by in vitro transcription, we placed the T7 promoter sequence at the 5’ end of the gRNA scaffold to form a 115nt-long oligo (gRNA-sca-temp, see Supplementary Table 1). We then designed 23-nt target sequences using an online program by selecting TTN as PAM sequence (http://www.rgenome.net/cas-designer/) [32]. The target sequences were extended at their 5’ end by adding a 19-nt sequence CAAATCTGAGAAGTGGCAC identical to the 3’ end of the gRNA scaffold. The reverse-compliment oligos of the extended target sequences (Table 1) were used together with a forward T7 promoter oligo (Table 1) and gRNA-sca-temp to generate double-stranded DNA by PCR reaction (with Q5 DNA polymerase, NEB, #M0491L). gRNA was synthesised using the PCR product as template with T7 RNA polymerase (Thermo Scientific, #EP0111) and template DNA was removed by DNaseI (Thermo Scientific, #EN0521) digestion. The gRNAs were further purified using 12% denaturing polyacrylamide gel.

### VarLOCK assay

Inspired by the potential for performing LAMP-SHERLOCK-coupled one-pot assay, we chose to perform the VarLOCK assay in Bst2.0 based buffer omitting dNTP and LAMP primers. For fluorescent assay, each 20 µl VarLOCK reaction contained 20 mM Tris-HCl pH 8.8@25°C, 10 mM (NH4)2SO4, 50 mM KCL, 8 mM MgSO4, 0.1% Tween 20, 37.5 nM AapCas12b, 12.5 nM gRNA, 200 nM fluorescent reporter and 10 nM double-stranded target DNA, unless stated otherwise. Fluorescent VarLOCK reaction were either performed at 60 or 65 °C (as indicated) and the FAM fluorescence was recorded every minute for 60 minutes with LightCycler 96 System (Roche) or ABI QuantStudio 7 (Thermo Fisher Scientific). For comparison, the initial reaction rates are calculated using linear regression in MS Excel by fitting the fluorescence increase during the first six minutes of the reaction. For lateral flow assay (LFA), all components were the same as above, except 375 nM AapCas12b, 125 nM gRNA, 125 nM LFA reporter and 100 nM double-stranded target DNA were used. LFA samples were incubated at 60 or 65 °C for 60 minutes and subsequently applied to dipsticks for result readout (Milenia HybriDetect 1, TwistDx, #MILENIA01) according to manufacturer’s instruction. Both fluorescent Reporter (5’-HEX-TTTTTTT-3’-IABkFQ/) and LFA reporter (5’-6-FAM-TTTTTTT-3’Bio) were purchased from IDT^4^ (REF#4Joung2020). To optimise the reaction, double-stranded short DNA, annealed using two complimentary oligos, were initially used as targets (sequence listed in Supplementary Table 2). For viral samples, RT-PCR products were generated using primers (listed in Supplementary Table 3) and used as targets in the VarLOCK assay.

### VarLOCK assay with PCR or LAMP products

Synthetic DNA templates harbouring the wt or mutant sequences for PCR or LAMP amplification were synthesised by IDT or by Twist Bioscience. Inactivated wt and mutant SARS-CoV-19 viral particles were provided by Dr. Richard Stanton and the viral RNA extracted with BOMB protocol [33]. LAMP primers were designed using the NEB LAMP primer design tool https://lamp.neb.com/#!/ or Primer Explorer v5.0 https://primerexplorer.jp/e/ (Eiken, Japan). qPCR primers were designed with IDT PrimerQuest Tool. Both LAMP and PCR primers were synthesised by IDT or Merck (listed in Supplementary Table S3). Luna Universal One-Step RT-qPCR Kit (NEB #E3005) was used for qPCR. When using viral RNA as template, 1x Luna WarmStart® RT Enzyme Mix was included and a 10 min incubation step at 50C was added before PCR cycles. For the LAMP reaction to produce amplicons for VarLOCK, the reaction mix contained 1X isothermal buffer (NEB), 10 mM DTT, 0.4 mg/ml PVP, 60 mM KCl, 5 mM each dNTP, 0.8 M betaine, 0.5 µM syto9, 0.32 U/µl Bst v2.0 Warm Start polymerase (NEB), 0.8 µM FIP/BIP, 0.4 µM LF/LB and 0.2 µM per 10 µl volume. Artificial templates were used for wild type, Alpha and Beta variants. LAMP amplification was observed on the green channel (490 nm) at 60 °C using a RotorGene thermo cycler (Qiagen) for 40 minutes.

### VarLOCK assay with saliva samples

Saliva samples were extracted in the Cardiff University COVID-19 test laboratory using automated BOMB protocol [33]. A pilot DNA quantification survey showed that typical concentration of the amplicons after RT-qPCR from saliva samples reached the range between 10 and 30 ng/µl, or a sub-picomole/µl in molar concentration, which is comfortably within the VarLOCK sensitivity range (1 to 100 femtomole/µl). Subsequently we did not quantify DNA concentration of the PCR products and directly used 2 µl for VarLOCK assay in 20 µl reaction. Optimised conditions were used for each of the mutation sites (Figure 5A).

### Collection and preparation of wastewater samples for VarLOCK assays

Untreated wastewater samples (crude influent) were collected between November 2020 and November 2021 from Dŵr Cymru/Welsh Water WwTWs at Cardiff, Newport and Swansea, as part of the Wales COVID-19 wastewater surveillance programme. On each sampling occasion, 500-1000 ml of crude influent was collected by manual grab sampling between 08.00 and 10.00 h, reflecting peak sewage flow and aiming to capture the highest community faecal load. Suspended solids were then removed from a 150 ml aliquot of each sample by centrifugation (3000 g, 4 °C, 30 min), and viral nucleic acids collected by subsequent PEG (polyethylene glycol 8000) precipitation for >18 hours based on methods described previously [34, 35] with some minor modifications [36]. PEG precipitates collected by centrifugation (10,000g, 4 °C, 30 min), and pellets, resuspended in 200 μl of phosphate buffer saline, prior to RNA extraction in the Cardiff University COVID-19 Testing Service laboratory using automated BOMB protocol [33].

### Automated liquid handling

The QPCR and reaction setup was automated on the Opentrons OT-2 robots. The OT-2 programs for assay assembly are available upon request.

### Sanger and Illumina sequencing

PCR products were sequenced using one of the two PCR primers by Eurofins Genomics sequencing service. Whole genome sequencing was performed utilising NGS library preparation by RT-PCR using the Nimagen protocol, essentially as described (https://www.protocols.io/view/wastewater-sequencing-using-the-easyseq-rc-pcr-sar-bx6dpra6). Subsequently, the libraries were reisolated with SPRI beads, quality of the libraries were checked with TapeStation and the libraries were quantified. Sequencing was performed on Illumina MiSeq 2×250 Nano cartridge.

### Statistics

Each experiment was performed with technical duplicates. Experiments with optimised condition were repeated between 2 and 5 times and Standard Deviation calculated from these biological replicates is presented in Figure 5B. Tests of the saliva samples were run with technical duplicates.

## Supporting information

Combined Supplementary Information

## Data Availability

All data produced in the present study are available upon reasonable request to the authors

## Acknowledgements

This work was supported by; UKRI/BBSRC emergency route for time-critical COVID-19 research, project number: BB/W003562/1 “Rapid Molecular Diagnostics For New & Emerging SARS-CoV-2 Variants of Concern-Protecting Vaccine Efficacy”, awarded to TPJ, OKC, JAHM, PH; Sêr Cymru Welsh Government award: MA/KW/1457/20 “Novel technologies for point-of-care genetic testing for SARS-CoV-2” awarded to TPJ, OKC, JAHM, PH; MRC (MR/V028448/1, MR/S00971X/1), Wellcome (204870/Z/16/Z), and Welsh Government Sêr Cymru calls awarded to RS. Wastewater project funded by Sêr Cymru awarded to PK and AW; Welsh Government “WeWASH” awarded to DJ, AW, TPJ, PK; Cardiff University funding in support of the establishment, implementation and operation of the CU COVID-19 screening service. We would like to thank Cardiff University Biobank for the assistance with sample collection and Dr. Angela Marchbank (School of Biosciences Genome Research Hub, Cardiff University) for help with Illumina sequencing. The authors would like to thank all users of Cardiff University COVID-19 screening service and all Cardiff University staff involved in its realisation.

## Author contributions

VarLOCK Conceptualization: XN, TPJ

Study concept, design, direction and development: XN, TPJ, OC, PH, PK, JAHM

Methodology: XN, DP, JG, JW, ZS, EH, GW, SH, PH, SD, JU, TPJ

Investigation: XN, SH, PH, SD, JU, DP, JG, JW, EH, GW, RS

Data analysis and visualization: XN, OC, PH, TPJ

Funding acquisition: OC, PH, JAHM, PK, DJ, AW, TPJ

Project administration: DJ, AW, TPJ

Supervision: JAHM, PH, OC, PK, AW, DJ, TPJ

Writing – original draft: TPJ, OC, XN, PH, JAHM

Writing – review & editing: All authors

## Ethical approval

I confirm all relevant ethical guidelines have been followed, and any necessary research ethics committee approvals have been obtained. The Ethical oversight is provided by Research Ethics Committee Health and Care Research Wales (IRAS Project ID: 214760). Saliva sample collection, processing and SARS-CoV-2 testing at the Cardiff University is covered by ethical approval 18/WA/0089 (Cardiff University BioBank). All necessary participant consent has been obtained and the appropriate institutional forms have been archived.

## Data availability statement

All biological materials used in this work can be obtained from Cardiff University Biobank on request. Data are available on request.

