## Supplementary material for "VarLOCK - sequencing independent, rapid detection of SARS-CoV-2 variants of concern for point-of-care testing, qPCR pipelines and national wastewater surveillance": Combined Supplementary Information


#### Authors list

Xinsheng Nan[1], Sven Hoehn[1], Patrick Hardinge[1], Shrinivas N Dighe[1], John Ukeri[2], Darius Pease[3], Joshua Griffin[3], Jessica I Warrington[1,5], Zack Saud[6], Emma Hottinger[3], Gordon Webster[1], Davey Jones[4], Peter Kille[1,3], Andrew Weightman[1], Richard Stanton[6], Oliver K Castell[2], James A.H. Murray[1], Tomasz P Jurkowski[1,3*]

#### Affiliation

[1]Cardiff School of Biosciences, Cardiff University, Sir Martin Evans Building, Museum Avenue, Cardiff, CF10 3AX, UK.
[2]Cardiff School of Pharmacy and Pharmaceutical Sciences, Cardiff University, Redwood Building, King Edward VII Avenue, Cardiff, CF10 3NB, UK.
[3]COVID-19 screening service, Cardiff University, Sir Martin Evans Building, Museum Avenue, Cardiff, CF10 3AX, UK.
[4]School of Natural Sciences, Bangor University, Bangor, Gwynedd, LL57 2UW, UK.
[5]Current address: Midatech Pharma (Wales) Ltd, 1 Caspian Point, Caspian Way, Cardiff, CF10 4DQ, UK.
[6] Infection & Immunity, School of Medicine, Cardiff University, Heath Park, Cardiff CF14 4XN, UK


### Supplementary materials

#### Figures


Supplementary Figure S1. Effects of chemical additives on detection sensitivities and specificities.
Supplementary Figure S2. VarLOCK specificity at different temperatures.
Supplementary Figure S3. VarLOCK reaction performed at 65°C with gRNA containing different lengths of target sequences in the presence of chemical additives.
Supplementary Figure S4. VarLOCK detection with PCR and LAMP products.
Supplementary Figure S5. Adaption of VarLOCK with lateral flow assay.
Supplementary Figure S6. Identification of the VOC with saliva samples collected at three periods of time.
Supplementary Figure S7. Sanger sequencing of selected mutations.


#### Tables


Supplementary Table S1. Sequences of guide RNAs.
Supplementary Table S2. Sequences of short target oligonucleotides.
Supplementary Table S3. Sequences of PCR and LAMP oligonucleotides.
Supplementary Table S4. Sequences of DNA synthetic templates for wild type and variants.
Supplementary Table S5. Chemical additives tested for reaction optimisation.

***Supplementary Figure S1. Effects of chemical additives on detection sensitivities and specificities.*** *Activities are shown as the rate of increase of arbitrary fluoresce (units/min).* ***A****, L18F/T20N site.* ***B*** *and* ***C****, E484K site.* ***D****, N501Y site. Wild type detection (wt>wt), wild type cross-reaction (wt>mut), mutation cross-reaction (mut>wt) and mutation detection (mut>mut).*

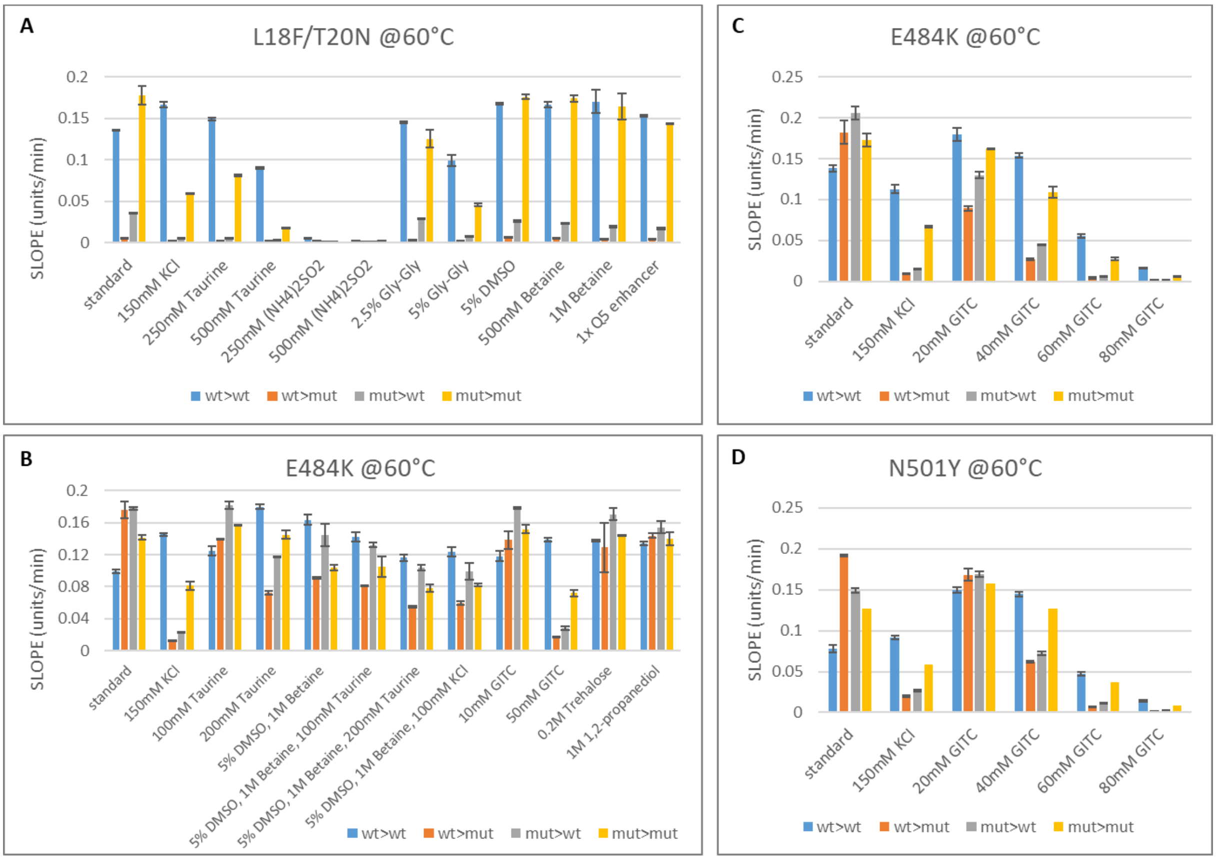

***Supplementary Figure S2. VarLOCK specificity at different temperatures.*** *VarLOCK reaction performed at 60°C (**A**, **C**, **E**, and **G**) and 65°C (**B**, **D**, **F** and **H**) with gRNA containing different lengths of target sequences. Wild type detection (wt>wt), wild type cross-reaction (wt>mut), mutation cross-reaction (mut>wt) and mutation detection (mut>mut).*

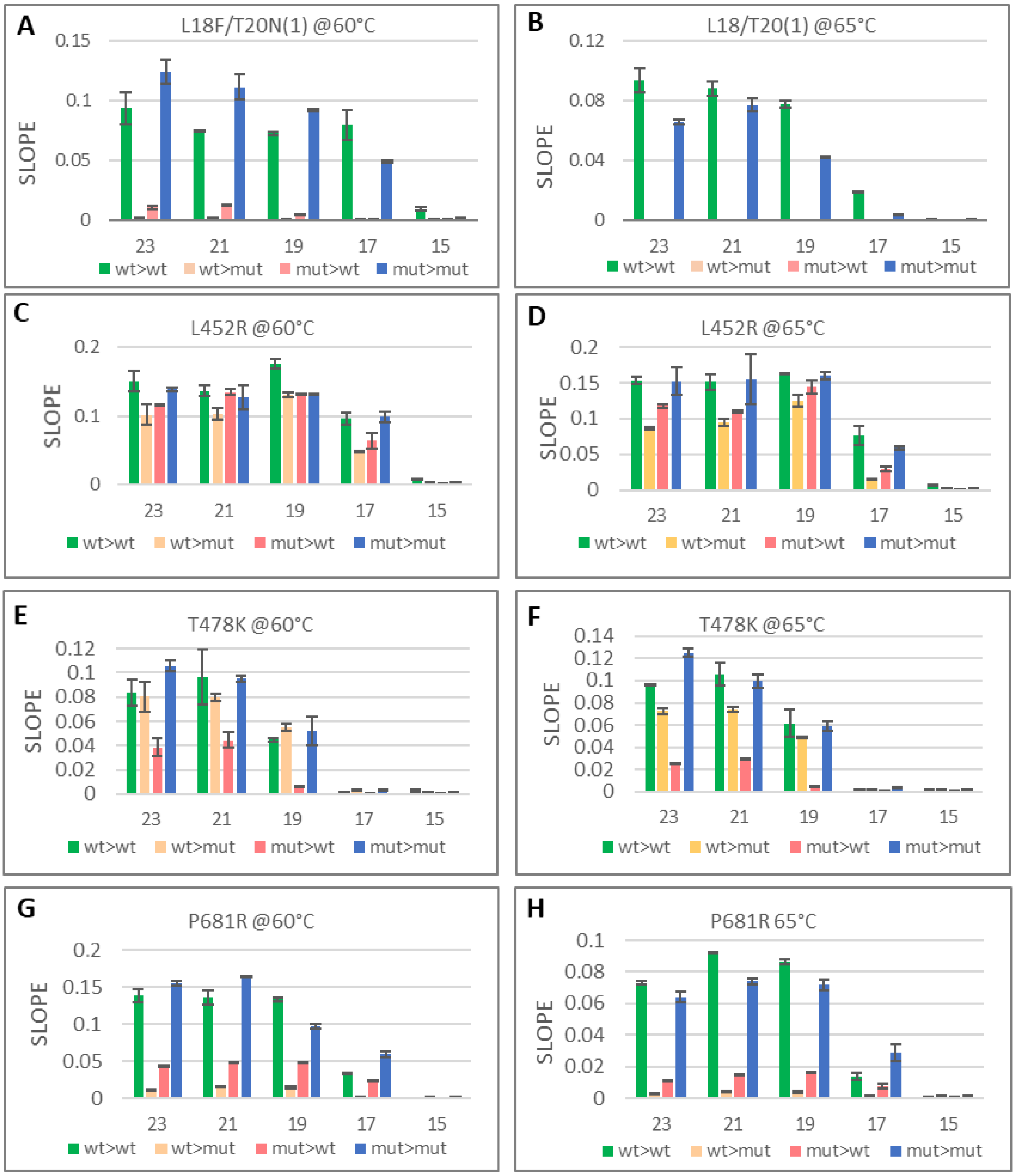

***Supplementary Figure S3. VarLOCK reaction performed at 65°C with gRNA containing different lengths of target sequences in the presence of chemical additives.*** *VarLOCK activities are plotted in panel **A** and **C**. Ratios of VarLOCK activities triggered by gRNA with matched target against mismatched target are shown in panel **B** and **D**. Wild type detection (wt>wt), wild type cross-reaction (wt>mut), mutation cross-reaction (mut>wt) and mutation detection (mut>mut).*

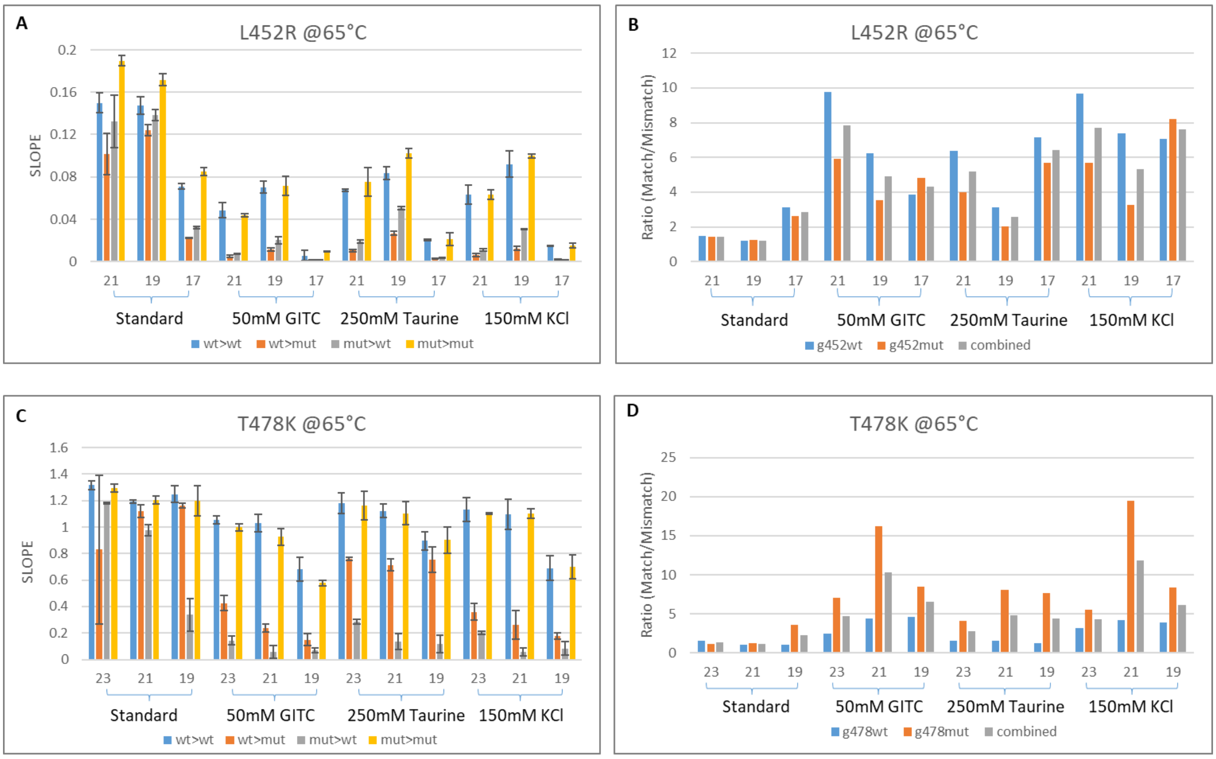

***Supplementary Figure S4. VarLOCK detection with PCR and LAMP products. A-D**, with PCR products as target for HV69/70del (**A**,**B**) and N501Y (**C**,**D**). **E-G**, with LAMP products as targets for HV69/70del (**E**), E484K (**F**) and N501Y (**G**).*

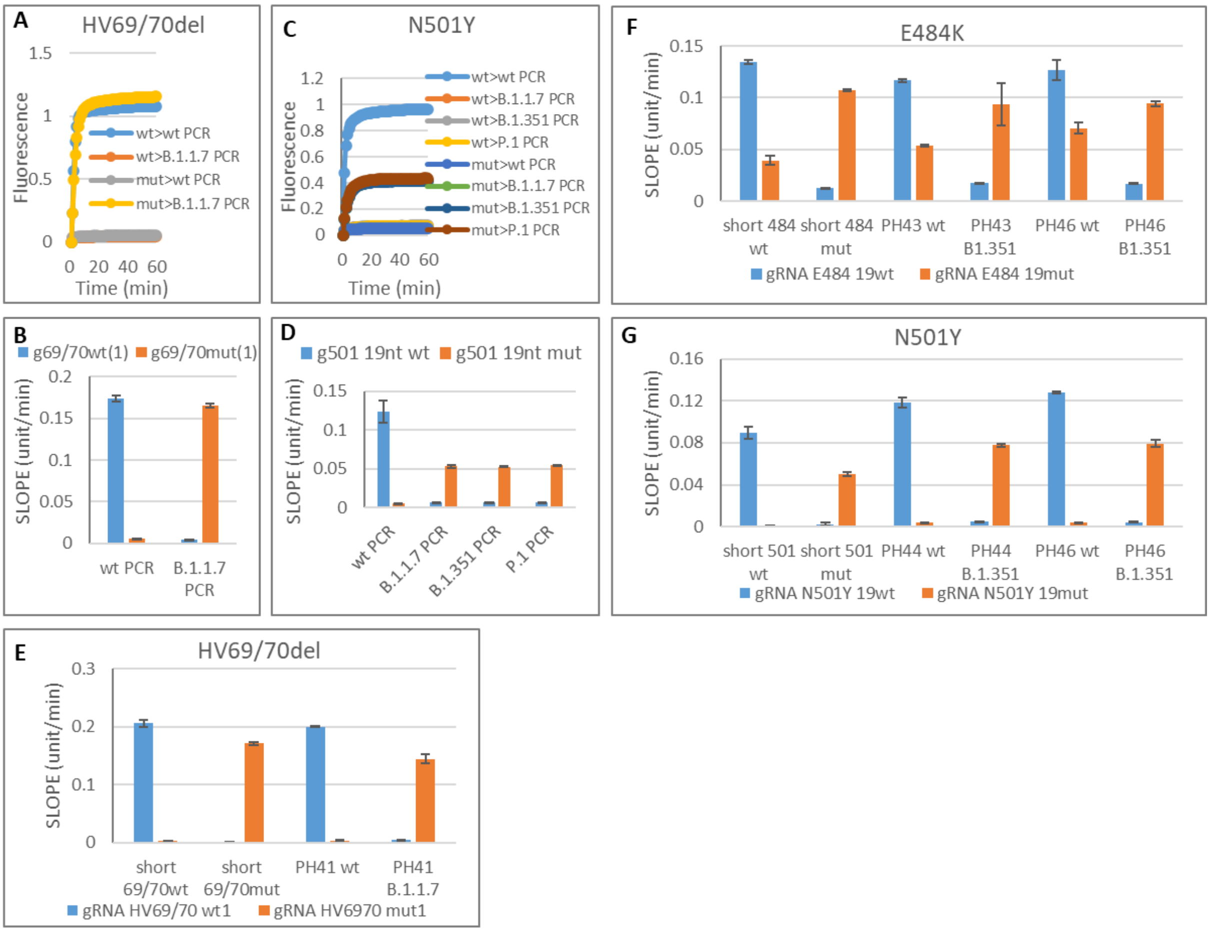

***Supplementary Figure S5. Adaptation of VarLOCK with lateral flow assay***. *gRNAs specific to wildtype or mutant sequence were incubated with wildtype (wt), mutant target (mut) or no target (N) in optimised condition (same as Fig 5C and I, except FAM-Biotin reporter was used instead of HEX-IABkFQ reporter). Endpoint reaction was applied to Milenia HybriDetect 1 Dipstick to visualise reactivity. Band intensity measured using ImageJ for relative comparison.*

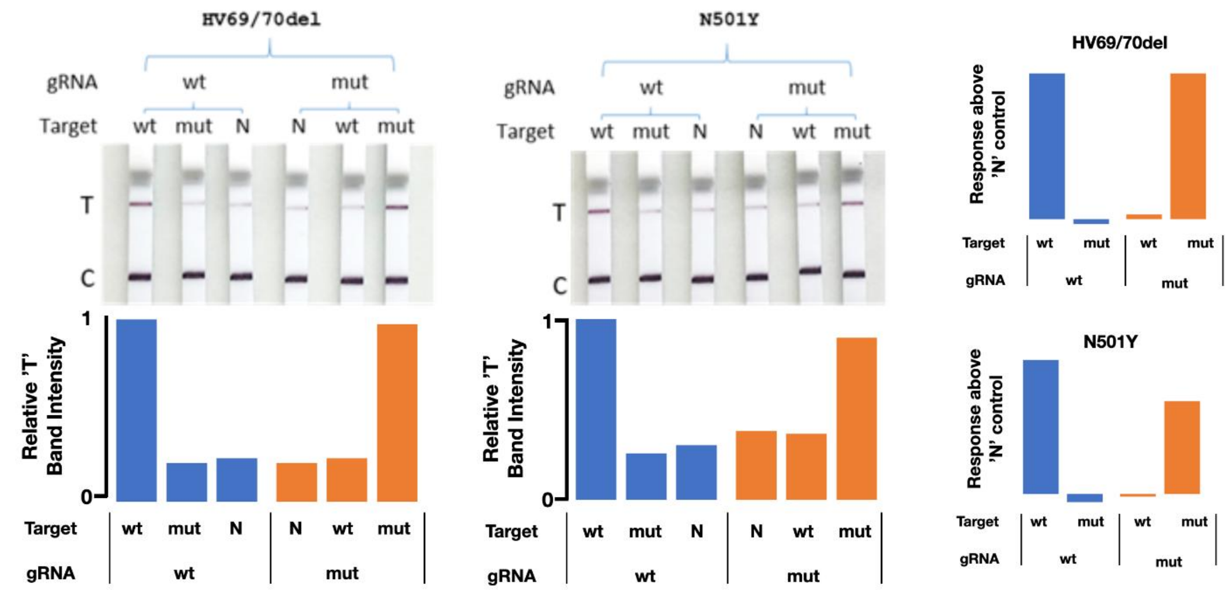

***Supplementary Figure S6. Identification of the VOC with saliva samples collected at three periods of time.*** *Firstly, the prior appearance of Alpha strain, the period when Alpha was in dominance and finally a period when Delta strain was in dominant.*

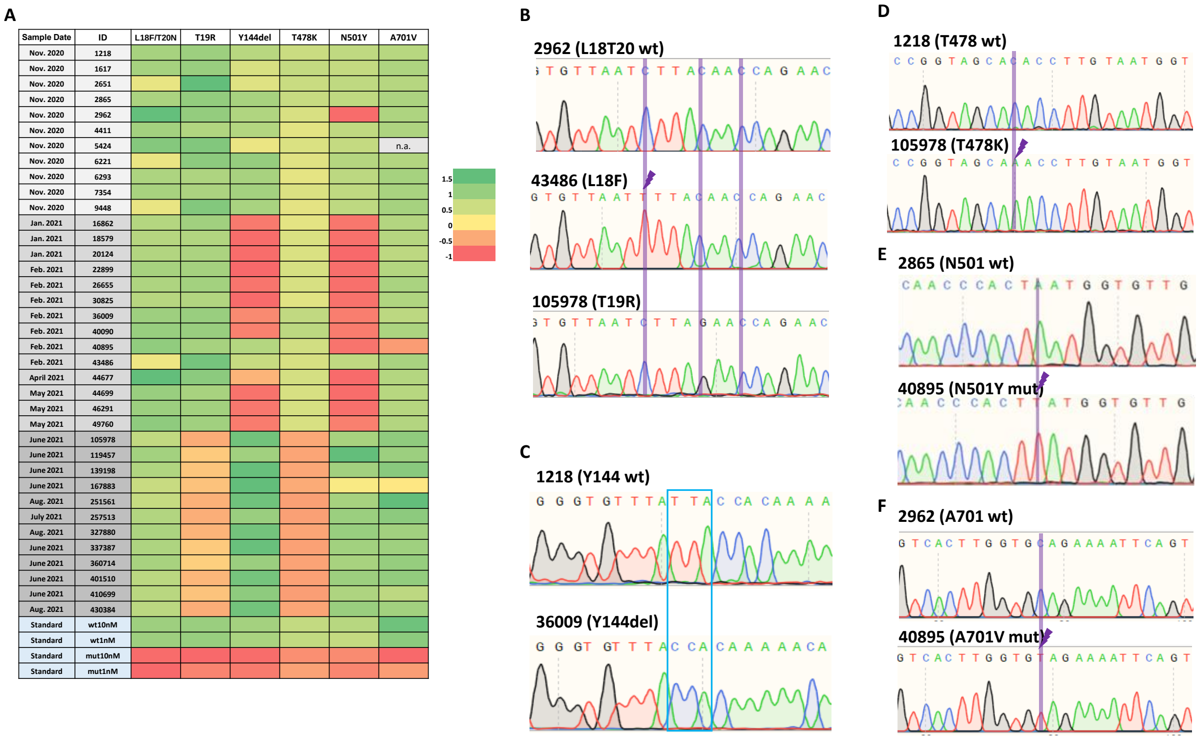

***Supplementary Figure S7. Sanger sequencing of selected mutations.*** *Sanger sequencing revealed that sample 167883 may contain a mixture of mutations that made the reading unrecognisable in some stretches, or that quality of the sample may be poor for sequencing, compared with a wildtype control sample 119457. A small proportion of A701V (C to T) mutation appeared to be present, indicated with vertical orange line. The affected codon is marked with box.*

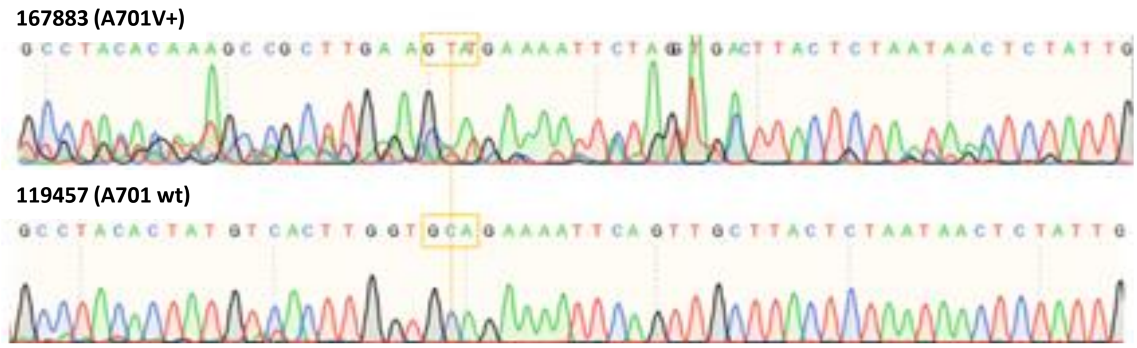

***Supplementary Table S1. Sequences of guide RNAs.*** *Nucleotides highlighted in red indicate the positions of substitutions and deletions.*

| Name | Sequence | Purpose | Reference |
|---|---|---|---|
| gRNA-sca-temp | gaaattaatacgactcactataggGTCTAGAGGACAGAATTTT TCAACGGGTGTGCCAATGGCCACTTTCCAGGTGGCAAAGCCCG TTGAGCTTCTCAAATCTGAGAAGTGGCAC | gRNA template | Joung et al |
| T7-primer-F | GAAATTAATACGACTCACTATAG | PCR primer for gRNA | this study |
| gRNA L18/T20 wt1-R | TAATTGAGTTCTGGTTGTAAGATGTGCCACTTCTCAGATTTG | PCR primer for gRNA | this study |
| gRNA L18F/T20N mut1-R | TAATTGAGTTCTGTTTGTAAAATGTGCCACTTCTCAGATTTG | PCR primer for gRNA | this study |
| gRNA L18/T20 wt2-R | TCAGTGTGTTAATCTTACAACCAGTGCCACTTCTCAGATTTG | PCR primer for gRNA | this study |
| gRNA L18F/T20N mut2-R | TCAGTGTGTTAATTTTACAAACAGTGCCACTTCTCAGATTTG | PCR primer for gRNA | this study |
| gRNA H69 wt1-R | CATGCTATACATGTCTCTGGGACGTGCCACTTCTCAGATTTG | PCR primer for gRNA | this study |
| gRNA H69 del1-R | TGGTTCCATGCTATCTCTGGGACGTGCCACTTCTCAGATTTG | PCR primer for gRNA | this study |
| gRNA H69 wt2-R | GTCCCAGAGACATGTATAGCATGGTGCCACTTCTCAGATTTG | PCR primer for gRNA | this study |
| gRNA H69 del2-R | ccattgGTCCCAGAGATAGCATGGTGCCACTTCTCAGATTTG | PCR primer for gRNA | this study |
| gRNA D80 wt-R | ATTAAATGGTAGGACAGGGTTATGTGCCACTTCTCAGATTTG | PCR primer for gRNA | this study |
| gRNA D80A mut-R | ATTAAATGGTAGGACAGGGTTAGGTGCCACTTCTCAGATTTG | PCR primer for gRNA | this study |
| gRNA Y144 wt-R | TGTTTTTGTGGTAATAAACACCCGTGCCACTTCTCAGATTTG | PCR primer for gRNA | this study |
| gRNA Y144 del-R | tgtTGTTTTTGTGGTAAACACCCGTGCCACTTCTCAGATTTG | PCR primer for gRNA | this study |
| gRNA E484 wt-R | CCTTGTAATGGTGTTGAAGGTTTGTGCCACTTCTCAGATTTG | PCR primer for gRNA | this study |
| gRNA E484K mut-R | CCTTGTAATGGTGTTAAAGGTTTGTGCCACTTCTCAGATTTG | PCR primer for gRNA | this study |
| gRNA N501 wt-R | TAACCAACACCATTAGTGGGTTGGTGCCACTTCTCAGATTTG | PCR primer for gRNA | this study |
| gRNA N501Y mut-R | TAACCAACACCATAAGTGGGTTGGTGCCACTTCTCAGATTTG | PCR primer for gRNA | this study |
| gRNA A701 wt-R | GTAAGCAACTGAATTTTCTGCACGTGCCACTTCTCAGATTTG | PCR primer for gRNA | this study |
| gRNA A701V mut-R | GTAAGCAACTGAATTTTCTACACGTGCCACTTCTCAGATTTG | PCR primer for gRNA | this study |
| gRNA A701 wt2-R | CCTACACTATGTCACTTGGTGCAGTGCCACTTCTCAGATTTG | PCR primer for gRNA | this study |
| gRNA A701V mut2-R | CCTACACTATGTCACTTGGTGTAGTGCCACTTCTCAGATTTG | PCR primer for gRNA | this study |
| gRNA L18/T20 21wt-R | ATTGAGTTCTGGTTGTAAGATGTGCCACTTCTCAGATTTG | PCR primer for gRNA | this study |
| gRNA L18/T20 19wt-R | TGAGTTCTGGTTGTAAGATGTGCCACTTCTCAGATTTG | PCR primer for gRNA | this study |
| gRNA L18/T20 17wt-R | AGTTCTGGTTGTAAGATGTGCCACTTCTCAGATTTG | PCR primer for gRNA | this study |
| gRNA L18/T20 15wt-R | TTCTGGTTGTAAGATGTGCCACTTCTCAGATTTG | PCR primer for gRNA | this study |
| gRNA L18F/T20N 21mut-R | ATTGAGTTCTGTTTGTAAAATGTGCCACTTCTCAGATTTG | PCR primer for gRNA | this study |
| gRNA L18F/T20N 19mut-R | TGAGTTCTGTTTGTAAAATGTGCCACTTCTCAGATTTG | PCR primer for gRNA | this study |
| gRNA L18F/T20N 17mut-R | AGTTCTGTTTGTAAAATGTGCCACTTCTCAGATTTG | PCR primer for gRNA | this study |
| gRNA L18F/T20N 15mut-R | TTCTGTTTGTAAAATGTGCCACTTCTCAGATTTG | PCR primer for gRNA | this study |
| gRNA E484 21wt-R | TTGTAATGGTGTTGAAGGTTTGTGCCACTTCTCAGATTTG | PCR primer for gRNA | this study |
| gRNA E484 19wt-R | GTAATGGTGTTGAAGGTTTGTGCCACTTCTCAGATTTG | PCR primer for gRNA | this study |
| gRNA E484 17wt-R | AATGGTGTTGAAGGTTTGTGCCACTTCTCAGATTTG | PCR primer for gRNA | this study |
| gRNA E484 15wt-R | TGGTGTTGAAGGTTTGTGCCACTTCTCAGATTTG | PCR primer for gRNA | this study |
| gRNA E484K 21mut-R | TTGTAATGGTGTTAAAGGTTTGTGCCACTTCTCAGATTTG | PCR primer for gRNA | this study |
| gRNA E484K 19mut-R | GTAATGGTGTTAAAGGTTTGTGCCACTTCTCAGATTTG | PCR primer for gRNA | this study |
| gRNA E484K 17mut-R | AATGGTGTTAAAGGTTTGTGCCACTTCTCAGATTTG | PCR primer for gRNA | this study |
| gRNA E484K 15mut-R | TGGTGTTAAAGGTTTGTGCCACTTCTCAGATTTG | PCR primer for gRNA | this study |
| gRNA N501Y 21wt-R | ACCAACACCATTAGTGGGTTGGTGCCACTTCTCAGATTTG | PCR primer for gRNA | this study |
| gRNA N501Y 19wt-R | CAACACCATTAGTGGGTTGGTGCCACTTCTCAGATTTG | PCR primer for gRNA | this study |
| gRNA N501Y 17wt-R | ACACCATTAGTGGGTTGGTGCCACTTCTCAGATTTG | PCR primer for gRNA | this study |
| gRNA N501Y 15wt-R | ACCATTAGTGGGTTGGTGCCACTTCTCAGATTTG | PCR primer for gRNA | this study |
| gRNA N501Y 21mut-R | ACCAACACCATAAGTGGGTTGGTGCCACTTCTCAGATTTG | PCR primer for gRNA | this study |
| gRNA N501Y 19mut-R | CAACACCATAAGTGGGTTGGTGCCACTTCTCAGATTTG | PCR primer for gRNA | this study |
| gRNA N501Y 17mut-R | ACACCATAAGTGGGTTGGTGCCACTTCTCAGATTTG | PCR primer for gRNA | this study |
| gRNA N501Y 15mut-R | ACCATAAGTGGGTTGGTGCCACTTCTCAGATTTG | PCR primer for gRNA | this study |

| | | | |
|---|---|---|---|
| gRNA N501Y 21wt-R | ACCAACACCATTAGTGGGTTGGTGCCACTTCTCAGATTTG | PCR primer for gRNA | this study |
| gRNA N501Y 19wt-R | CAACACCATTAGTGGGTTGGTGCCACTTCTCAGATTTG | PCR primer for gRNA | this study |
| gRNA N501Y 17wt-R | ACACCATTAGTGGGTTGGTGCCACTTCTCAGATTTG | PCR primer for gRNA | this study |
| gRNA N501Y 15wt-R | ACCATTAGTGGGTTGGTGCCACTTCTCAGATTTG | PCR primer for gRNA | this study |
| gRNA N501Y 21mut-R | ACCAACACCATAAGTGGGTTGGTGCCACTTCTCAGATTTG | PCR primer for gRNA | this study |
| gRNA N501Y 19mut-R | CAACACCATAAGTGGGTTGGTGCCACTTCTCAGATTTG | PCR primer for gRNA | this study |
| gRNA N501Y 17mut-R | ACACCATAAGTGGGTTGGTGCCACTTCTCAGATTTG | PCR primer for gRNA | this study |
| gRNA N501Y 15mut-R | ACCATAAGTGGGTTGGTGCCACTTCTCAGATTTG | PCR primer for gRNA | this study |
| gRNA L452wt-R23 | ACTTCCTAAACAATCTATACAGGGTGCCACTTCTCAGATTTG | PCR primer for gRNA | this study |
| gRNA L452wt-R21 | TTCCTAAACAATCTATACAGGGTGCCACTTCTCAGATTTG | PCR primer for gRNA | this study |
| gRNA L452wt-R19 | CCTAAACAATCTATACAGGGTGCCACTTCTCAGATTTG | PCR primer for gRNA | this study |
| gRNA L452wt-R17 | TAAACAATCTATACAGGGTGCCACTTCTCAGATTTG | PCR primer for gRNA | this study |
| gRNA L452wt-R15 | AACAATCTATACAGGGTGCCACTTCTCAGATTTG | PCR primer for gRNA | this study |
| gRNA L452R-R23 | ACTTCCTAAACAATCTATACCGGGTGCCACTTCTCAGATTTG | PCR primer for gRNA | this study |
| gRNA L452R-R21 | TTCCTAAACAATCTATACCGGGTGCCACTTCTCAGATTTG | PCR primer for gRNA | this study |
| gRNA L452R-R19 | CCTAAACAATCTATACCGGGTGCCACTTCTCAGATTTG | PCR primer for gRNA | this study |
| gRNA L452R-R17 | TAAACAATCTATACCGGGTGCCACTTCTCAGATTTG | PCR primer for gRNA | this study |
| gRNA L452R-R15 | AACAATCTATACCGGGTGCCACTTCTCAGATTTG | PCR primer for gRNA | this study |
| gRNA T478wt-R23 | TATCAGGCCGGTAGCACACCTTGGTGCCACTTCTCAGATTTG | PCR primer for gRNA | this study |
| gRNA T478wt-R21 | TCAGGCCGGTAGCACACCTTGGTGCCACTTCTCAGATTTG | PCR primer for gRNA | this study |
| gRNA T478wt-R19 | AGGCCGGTAGCACACCTTGGTGCCACTTCTCAGATTTG | PCR primer for gRNA | this study |
| gRNA T478wt-R17 | GCCGGTAGCACACCTTGGTGCCACTTCTCAGATTTG | PCR primer for gRNA | this study |
| gRNA T478wt-R15 | CGGTAGCACACCTTGGTGCCACTTCTCAGATTTG | PCR primer for gRNA | this study |
| gRNA T478K-R23 | TATCAGGCCGGTAGCAAACCTTGGTGCCACTTCTCAGATTTG | PCR primer for gRNA | this study |
| gRNA T478K-R21 | TCAGGCCGGTAGCAAACCTTGGTGCCACTTCTCAGATTTG | PCR primer for gRNA | this study |
| gRNA T478K-R19 | AGGCCGGTAGCAAACCTTGGTGCCACTTCTCAGATTTG | PCR primer for gRNA | this study |
| gRNA T478K-R17 | GCCGGTAGCAAACCTTGGTGCCACTTCTCAGATTTG | PCR primer for gRNA | this study |
| gRNA T478K-R15 | CGGTAGCAAACCTTGGTGCCACTTCTCAGATTTG | PCR primer for gRNA | this study |
| gRNA P681wt-R23 | CTACACTACGTGCCCGCCGAGGAGTGCCACTTCTCAGATTTG | PCR primer for gRNA | this study |
| gRNA P681wt-R21 | ACACTACGTGCCCGCCGAGGAGTGCCACTTCTCAGATTTG | PCR primer for gRNA | this study |
| gRNA P681wt-R19 | ACTACGTGCCCGCCGAGGAGTGCCACTTCTCAGATTTG | PCR primer for gRNA | this study |
| gRNA P681wt-R17 | TACGTGCCCGCCGAGGAGTGCCACTTCTCAGATTTG | PCR primer for gRNA | this study |
| gRNA P681wt-R15 | CGTGCCCGCCGAGGAGTGCCACTTCTCAGATTTG | PCR primer for gRNA | this study |
| gRNA P681R-R23 | CTACACTACGTGCCCGCCGACGAGTGCCACTTCTCAGATTTG | PCR primer for gRNA | this study |
| gRNA P681R-R21 | ACACTACGTGCCCGCCGACGAGTGCCACTTCTCAGATTTG | PCR primer for gRNA | this study |
| gRNA P681R-R19 | ACTACGTGCCCGCCGACGAGTGCCACTTCTCAGATTTG | PCR primer for gRNA | this study |
| gRNA P681R-R17 | TACGTGCCCGCCGACGAGTGCCACTTCTCAGATTTG | PCR primer for gRNA | this study |
| gRNA P681R-R15 | CGTGCCCGCCGACGAGTGCCACTTCTCAGATTTG | PCR primer for gRNA | this study |
| gRNA T19R-R23 | TAATTGAGTTCTGGTTCTAAGATGTGCCACTTCTCAGATTTG | PCR primer for gRNA | this study |
| gRNA T19R-R21 | ATTGAGTTCTGGTTCTAAGATGTGCCACTTCTCAGATTTG | PCR primer for gRNA | this study |
| gRNA T19R-R19 | TGAGTTCTGGTTCTAAGATGTGCCACTTCTCAGATTTG | PCR primer for gRNA | this study |
| gRNA T19R-R17 | AGTTCTGGTTCTAAGATGTGCCACTTCTCAGATTTG | PCR primer for gRNA | this study |
| gRNA T19R-R15 | TTCTGGTTCTAAGATGTGCCACTTCTCAGATTTG | PCR primer for gRNA | this study |

***Supplementary Table S2. Sequences of short target oligonucleotides.*** *Nucleotides highlighted in red indicate the positions of substitutions and deletions.*

| Name | Sequence | Purpose | Reference |
|---|---|---|---|
| Temp L18/T20 wt-F | CAGTGTGTTAATCTTACAACCAGAACTCAATTACCC | gRNA target | this study |
| Temp L18/T20 wt-R | GGGTAATTGAGTTCTGGTTGTAAGATTAACACACTG | gRNA target | this study |
| Temp L18F/T20N mut-F | CAGTGTGTTAATTTTACAAACAGAACTCAATTACCC | gRNA target | this study |
| Temp L18F/T20N mut-R | GGGTAATTGAGTTCTGTTTGTAAAATTAACACACTG | gRNA target | this study |
| Temp L18/T20 wt2-F | TCTAGTCAGTGTGTTAATCTTACAACCAGAACTCAA | gRNA target | this study |
| Temp L18/T20 wt2-R | TTGAGTTCTGGTTGTAAGATTAACACACTGACTAGA | gRNA target | this study |
| Temp L18F/T20N mut2-F | TCTAGTCAGTGTGTTAATTTTACAAACAGAACTCAA | gRNA target | this study |
| Temp L18F/T20N mut2-R | TTGAGTTCTGTTTGTAAAATTAACACACTGACTAGA | gRNA target | this study |
| Temp H69 wt-F | ACTTGGTTCCATGCTATACATGTCTCTGGGACCAATGGTAC | gRNA target | this study |
| Temp H69 wt-R | GTACCATTGGTCCCAGAGACATGTATAGCATGGAACCAAGT | gRNA target | this study |
| Temp H69 del-F | ACTTGGTTCCATGCTATCTCTGGGACCAATGGTACTAAG | gRNA target | this study |
| Temp H69 del-R | CTTAGTACCATTGGTCCCAGAGATAGCATGGAACCAAGT | gRNA target | this study |
| Temp D80 wt-F | AAGAGGTTTGATAACCCTGTCCTACCATTTAATGATG | gRNA target | this study |
| Temp D80 wt-R | CATCATTAAATGGTAGGACAGGGTTATCAAACCTCTT | gRNA target | this study |
| Temp D80A mut-F | AAGAGGTTTGCTAACCCTGTCCTACCATTTAATGATG | gRNA target | this study |
| Temp D80A mut-R | CATCATTAAATGGTAGGACAGGGTTAGCAAACCTCTT | gRNA target | this study |
| Temp Y144 wt-F | ATCCATTTTTGGGTGTTTATTACCACAAAAACAACA | gRNA target | this study |
| Temp Y144 wt-R | TGTTGTTTTTGTGGTAATAAACACCCAAAAATGGAT | gRNA target | this study |
| Temp Y144 del-F | ATCCATTTTTGGGTGTTTACCACAAAAACAACAAAA | gRNA target | this study |
| Temp Y144 del-R | TTTTGTTGTTTTTGTGGTAAACACCCAAAAATGGAT | gRNA target | this study |
| Temp E484 wt-F | CACACCTTGTAATGGTGTTGAAGGTTTTAATTGTTA | gRNA target | this study |
| Temp E484 wt-R | TAACAATTAAAACCTTCAACACCATTACAAGGTGTG | gRNA target | this study |
| Temp E484K mut-F | CACACCTTGTAATGGTGTTAAAGGTTTTAATTGTTA | gRNA target | this study |
| Temp E484K mut-R | TAACAATTAAAACCTTTAACACCATTACAAGGTGTG | gRNA target | this study |
| Temp N501 wt-F | TATGGTTTCCAACCCACTAATGGTGTTGGTTACCAA | gRNA target | this study |
| Temp N501 wt-R | TTGGTAACCAACACCATTAGTGGGTTGGAAACCATA | gRNA target | this study |
| Temp N501Y mut-F | TATGGTTTCCAACCCACTTATGGTGTTGGTTACCAA | gRNA target | this study |
| Temp N501Y mut-R | TTGGTAACCAACACCATAAGTGGGTTGGAAACCATA | gRNA target | this study |
| Temp A701 wt-F | GTCACTTGGTGCAGAAAATTCAGTTGCTTACTCTAA | gRNA target | this study |
| Temp A701 wt-R | TTAGAGTAAGCAACTGAATTTTCTGCACCAAGTGAC | gRNA target | this study |
| Temp A701V mut-F | GTCACTTGGTGTAGAAAATTCAGTTGCTTACTCTAA | gRNA target | this study |
| Temp A701V mut-R | TTAGAGTAAGCAACTGAATTTTCTACACCAAGTGAC | gRNA target | this study |
| Temp A701 wt2-F | ATCATTGCCTACACTATGTCACTTGGTGCAGAAAATT | gRNA target | this study |
| Temp A701 wt2-R | AATTTTCTGCACCAAGTGACATAGTGTAGGCAATGAT | gRNA target | this study |
| Temp A701V2 mut-F | ATCATTGCCTACACTATGTCACTTGGTGTAGAAAATT | gRNA target | this study |
| Temp A701V2 mut-R | AATTTTCTACACCAAGTGACATAGTGTAGGCAATGAT | gRNA target | this study |

*Supplementary Table S3. Sequences of PCR and LAMP oligonucleotides.*

| Oligo Name | Sequence | F primer start | R primer end | PCR product length (wt) | Reference |
|---|---|---|---|---|---|
| PH40 L18T20 F3 | CAACAGAGTTGTTATTTCTAGTGAT | | | | |
| PH40 L18T20 B3 | ACAAGTCCTGAGTTGAATGT | 21514 | 21725 | 212 | this study |
| 69/70-F | TCAACTCAGGACTTGTTCTTACCT | | | | |
| 69/70-R | TGGTAGGACAGGGTTATCAAAC | 21710 | 21817 | 108 | this study |
| 144-F | ACGCTACTAATGTTGTTATTAAAGTCT | | | | |
| 144-R | TCTGAACTCACTTTCCATCCAACT | 21927 | 22036 | 110 | this study |
| PH43 E484K F3 | GTTGGTGGTAATTATAATTACCTGT | | | | |
| PH43 E484K B3 | TGGTGCATGTAGAAGTTCAA | 22895 | 23125 | 231 | this study |
| PH46 484/501 F3 | TTGGTGGTAATTATAATTACCTGTA | | | | |
| PH46 484/501 B3 | TGACACATTTGTTTTTAACCAAAT | 22896 | 23180 | 285 | this study |
| PH44 N501Y F3 | TCTCAAACCTTTTGAGAGAGA | | | | |
| PH44 N501Y B3 | TTGACACATTTGTTTTTAACCAA | 22942 | 23181 | 240 | this study |
| PH51 A701V F3 | GACATACCCATTGGTGCA | | | | |
| PH51 A701V B3 | ATCTACTGATGTCTTGGTCAT | 23549 | 23773 | 225 | this study |

| Oligo Name | Sequence | F3 primer start | B3 primer end | LAMP coverage length (wt) | Reference |
|---|---|---|---|---|---|
| PH40 L18T20 F3 | CAACAGAGTTGTTATTTCTAGTGAT | | | | |
| PH40 L18T20 B3 | ACAAGTCCTGAGTTGAATGT | | | | |
| PH40 L18T20 FIP | ACTGACTAGAGACTAGTGGCAATAAGTTCTTGTTAACAACTAAACGAA | | | | |
| PH40 L18T20 BIP | CTGCATACACTAATTCTTTCACACGAACTGAGGATCTGAAAACTTTG | | | | |
| PH40 L18T20 LF | AACAAGAAAAACAAACATTG | | | | |
| PH40 L18T20 LB | TGGTGTTTATTACCCTGA | 21514 | 21725 | 212 | this study |
| PH41 69del70 F3 | TCTTTCACACGTGGTGTT | | | | |
| PH41 69del70 B3 | GTACCAAAAATCCAGCCTC | | | | |
| PH41 69del70 FIP | CAAGTAACATTGGAAAAGAAAGGTATACCCTGACAAAGTTTTCAG | | | | |
| PH41 69del70 BIP | CTAAGAGGTTTGATAACCCTGTCACTTCTCAGTGGAAGCAA | | | | |
| PH41 69del70 LF | AGTTGAATGTAAAACTGAGGAT | | | | |
| PH41 69del70 LB | CTACCATTTAATGATGGTGTTTATT | 21653 | 21885 | 233 | this study |
| PH42 Y144 F3 | GTACTACTTTAGATTCGAAGACC | | | | |
| PH42 Y144 B3 | CCTGTTTTCCTTCAAGGTCC | | | | |
| PH42 Y144 FIP | GGATCATTACAAAATTGAAATTCACCAGTCCCTACTTATTGTTAATAACG | | | | |
| PH42 Y144 BIP | GTTGGATGGAAAGTGAGTTCGAGAGACATATTCAAAAGTGCA | | | | |
| PH42 Y144 LF | AGACTTTAATAACAACATTAGTAG | | | | |
| PH42 Y144 LB | AGAGTTTATTCTAGTGCGAATAA | 21882 | 22112 | 231 | this study |
| PH43 E484K F3 | GTTGGTGGTAATTATAATTACCTGT | | | | |
| PH43 E484K B3 | TGGTGCATGTAGAAGTTCAA | | | | |
| PH43 E484K FIP | TGCTACCGGCCTGATAGATTTCTGTTTAGGAAGTCTAATCTCAAAC | | | | |
| PH43 E484K BIP | TACAATCATATGGTTTCCAACCCAAGAAAGTACTACTACTCTGTATGG | | | | |
| PH43 E484K LF | GTTGAAATATCTCTCTCAAAAG | | | | |
| PH43 E484K LB | CTAATGGTGTTGGTTACC | 22895 | 23125 | 231 | this study |
| PH44 N501Y F3 | TCTCAAACCTTTTGAGAGAGA | | | | |
| PH44 N501Y B3 | TTGACACATTTGTTTTTAACCAA | | | | |
| PH44 N501Y FIP | CCATATGATTGTAAAGGAAAGTAACCAACTGAAATCTATCAGGCC | | | | |
| PH44 N501Y BIP | CCATACAGAGTAGTAGTACTTTCTTTTTTAGGTCCACAAACAGT | | | | |
| PH44 N501Y LF | CACCATTACAAGGTGTGCTACC | | | | |
| PH44 N501Y LB | TTTGAACTTCTACATGCACCAGC | 22942 | 23181 | 240 | this study |
| PH46 484/501 F3 | TTGGTGGTAATTATAATTACCTGTA | | | | |
| PH46 484/501 B3 | TGACACATTTGTTTTTAACCAAAT | | | | |
| PH46 484/501 FIP | TGCTACCGGCCTGATAGATTGTTTAGGAAGTCTAATCTCAAACC | | | | |
| PH46 484/501 BIP | TTCTTTTGAACTTCTACATGCACCATTAGGTCCACAAACAGTTGC | | | | |
| PH46 484/501 LF | TCAGTTGAAATATCTCTCTCAAAA | 22896 | 23180 | 285 | this study |
| PH51 A701V F3 | GACATACCCATTGGTGCA | | | | |
| PH51 A701V B3 | ATCTACTGATGTCTTGGTCAT | | | | |
| PH51 A701V FIP | TGGATTGACTAGCTACACTACGTGTATATGCGCTAGTTATCAGACTC | | | | |
| PH51 A701V BIP | TAACTCTATTGCCATACCCACAAATACTGGTAGAATTTCTGTGGTAAC | | | | |
| PH51 A701V LF | CCCGCCGAGGAGAATTAGTCT | 23549 | 23773 | 225 | this study |

***Supplementary Table S4. Sequences of DNA synthetic templates for wild type and variants.***
*Nucleotides highlighted in red indicate the positions of substitutions and deletions.*

| Template Name | Sequence | Start position | Finish position | Template length | Reference |
|---|---|---|---|---|---|
| PHS1-4 (wt) | GTCAGTGTGTTAATCTTACAACCAGAACTCAATTACCCCCTGCATACACTAATTCT<br>TTCACACGTGGTGTTTATTACCCTGACAAAGTTTTCAGATCCTCAGTTTTACATTC<br>AACTCAGGACTTGTTCTTACCTTTCTTTTCCAATGTTACTTGGTTCCATGCTATACA<br>TGTCTCTGGGACCAATGGTACTAAGAGGTTTGATAACCCTGTCCTACCATTTAAT<br>GATGGTGTTTATTTTGCTTCCACTGAGAAGTCTAACATAATAAGAGGCTGGATTTT<br>TGGTACTACTTTAGATTCGAAGACCCAGTCCCTACTTATTGTTAATAACGCTACTA<br>ATGTTGTTATTAAAGTCTGTGAATT | 21600 | 21960 | 361 | this study |
| PHs1-4 (Alpha) | GTCAGTGTGTTAATCTTACAACCAGAACTCAATTACCCCCTGCATACACTAATTCT<br>TTCACACGTGGTGTTTATTACCCTGACAAAGTTTTCAGATCCTCAGTTTTACATTC<br>AACTCAGGACTTGTTCTTACCTTTCTTTTCCAATGTTACTTGGTTCCATGCTATacat<br>gtCTCTGGGACCAATGGTACTAAGAGGTTTGATAACCCTGTCCTACCATTTAATGA<br>TGGTGTTTATTTTGCTTCCACTGAGAAGTCTAACATAATAAGAGGCTGGATTTTTG<br>GTACTACTTTAGATTCGAAGACCCAGTCCCTACTTATTGTTAATAACGCTACTAAT<br>GTTGTTATTAAAGTCTGTGAATT | 21600 | 21960 | 355 | this study |
| 417 484 501 (wt) | GGAACAGGAAGAGAATCAGCAACTGTGTTGCTGATTATTCTGTCCTATATAATTC<br>CGCATCATTTTCCACTTTTAAGTGTTATGGAGTGTCTCCTACTAAATTAAATGATC<br>TCTGCTTTACTAATGTCTATGCAGATTCATTTGTAATTAGAGGTGATGAAGTCAGA<br>CAAATCGCTCCAGGGCAAACTGGAAAGATTGCTGATTATAATTATAAATTACCA<br>GATGATTTTACAGGCTGCGTTATAGCTTGGAATTCTAACAATCTTGATTCTAAGGT<br>TGGTGGTAATTATAATTACCTGTATAGATTGTTTAGGAAGTCTAATCTCAAACCTT<br>TTGAGAGAGATATTTCAACTGAAATCTATCAGGCCGGTAGCACACCTTGTAATGG<br>TGTTGAAGGTTTTAATTGTTACTTTCCTTTACAATCATATGGTTTCCAACCCACTAA<br>TGGTGTTGGTTACCAACCATACAGAGTAGTAGTACTTTCTTTTGAACTTCTACATG<br>CACCAGCAACTGTTTGTGGACCTAAAAAGTCTACTAATTTGGTTAAAAACAAATG<br>TGTCAATTTCAACTTCAATGGTTTAACAGGCACAGGTGTTCTTACTGAGTCTAAC | 22620 | 23230 | 610 | this study |
| 417 484 501 (Alpha) | GGAACAGGAAGAGAATCAGCAACTGTGTTGCTGATTATTCTGTCCTATATAATTC<br>CGCATCATTTTCCACTTTTAAGTGTTATGGAGTGTCTCCTACTAAATTAAATGATC<br>TCTGCTTTACTAATGTCTATGCAGATTCATTTGTAATTAGAGGTGATGAAGTCAGA<br>CAAATCGCTCCAGGGCAAACTGGAAAGATTGCTGATTATAATTATAAATTACCA<br>GATGATTTTACAGGCTGCGTTATAGCTTGGAATTCTAACAATCTTGATTCTAAGGT<br>TGGTGGTAATTATAATTACCTGTATAGATTGTTTAGGAAGTCTAATCTCAAACCTT<br>TTGAGAGAGATATTTCAACTGAAATCTATCAGGCCGGTAGCACACCTTGTAATGG<br>TGTTGAAGGTTTTAATTGTTACTTTCCTTTACAATCATATGGTTTCCAACCCACTTA<br>TGGTGTTGGTTACCAACCATACAGAGTAGTAGTACTTTCTTTTGAACTTCTACATG<br>CACCAGCAACTGTTTGTGGACCTAAAAAGTCTACTAATTTGGTTAAAAACAAATG<br>TGTCAATTTCAACTTCAATGGTTTAACAGGCACAGGTGTTCTTACTGAGTCTAAC | 22620 | 23230 | 610 | this study |
| 417 484 501 (Beta) | GGAACAGGAAGAGAATCAGCAACTGTGTTGCTGATTATTCTGTCCTATATAATTC<br>CGCATCATTTTCCACTTTTAAGTGTTATGGAGTGTCTCCTACTAAATTAAATGATC<br>TCTGCTTTACTAATGTCTATGCAGATTCATTTGTAATTAGAGGTGATGAAGTCAGA<br>CAAATCGCTCCAGGGCAAACTGGAAATATTGCTGATTATAATTATAAATTACCAG<br>ATGATTTTACAGGCTGCGTTATAGCTTGGAATTCTAACAATCTTGATTCTAAGGTT<br>GGTGGTAATTATAATTACCTGTATAGATTGTTTAGGAAGTCTAATCTCAAACCTTT<br>TGAGAGAGATATTTCAACTGAAATCTATCAGGCCGGTAGCACACCTTGTAATGGT<br>GTTAAAGGTTTTAATTGTTACTTTCCTTTACAATCATATGGTTTCCAACCCACTTAT<br>GGTGTTGGTTACCAACCATACAGAGTAGTAGTACTTTCTTTTGAACTTCTACATGC<br>ACCAGCAACTGTTTGTGGACCTAAAAAGTCTACTAATTTGGTTAAAAACAAATGT<br>GTCAATTTCAACTTCAATGGTTTAACAGGCACAGGTGTTCTTACTGAGTCTAAC | 22620 | 23230 | 610 | this study |
| 417 484 501 (Gamma) | GGAACAGGAAGAGAATCAGCAACTGTGTTGCTGATTATTCTGTCCTATATAATTC<br>CGCATCATTTTCCACTTTTAAGTGTTATGGAGTGTCTCCTACTAAATTAAATGATC<br>TCTGCTTTACTAATGTCTATGCAGATTCATTTGTAATTAGAGGTGATGAAGTCAGA<br>CAAATCGCTCCAGGGCAAACTGGAACGATTGCTGATTATAATTATAAATTACCA<br>GATGATTTTACAGGCTGCGTTATAGCTTGGAATTCTAACAATCTTGATTCTAAGGT<br>TGGTGGTAATTATAATTACCTGTATAGATTGTTTAGGAAGTCTAATCTCAAACCTT<br>TTGAGAGAGATATTTCAACTGAAATCTATCAGGCCGGTAGCACACCTTGTAATGG<br>TGTTAAAGGTTTTAATTGTTACTTTCCTTTACAATCATATGGTTTCCAACCCACTTA<br>TGGTGTTGGTTACCAACCATACAGAGTAGTAGTACTTTCTTTTGAACTTCTACATG<br>CACCAGCAACTGTTTGTGGACCTAAAAAGTCTACTAATTTGGTTAAAAACAAATG<br>TGTCAATTTCAACTTCAATGGTTTAACAGGCACAGGTGTTCTTACTGAGTCTAAC | 22620 | 23230 | 610 | this study |

*Supplementary Table S5. Chemical additives tested for reaction optimisation.*

| Chemicals | Range tested | Effects | Optimal concentraction |
|---|---|---|---|
| KCl | 50-200 mM | substantial | 150 mM |
| GITC | 20-60 mM | substantial | 50 mM |
| Taurine | 100-500 mM | moderate | 250 mM |
| Betaine | 0.5-1 M | moderate | 1 M |
| Q5 enhancer | 1x | small | 1x |
| $(NH_4)_2SO_4$ | 250-500 mM | inactivation | |
| Gly-Gly | 2.5-5 % | none | |
| DMSO | 5% | none | |
| Trehalose | 0.2 M | none | |
| 1,2-propanediol | 1 M | none | |
